# Subphenotypes of Self-Reported Symptoms and Outcomes in Long COVID: a prospective cohort study with latent class analysis

**DOI:** 10.1101/2023.08.09.23293901

**Authors:** Georgios D. Kitsios, Shawna Blacka, Jana Jacobs, Taaha Mirza, Asma Naqvi, Heather Gentry, Cathy Kessinger, Xiaohong Wang, Konstantin Golubykh, Hafiz Muhammad Siddique Qurashi, Akash Dodia, Michael Risbano, Michael Benigno, Birol Emir, Edward Weinstein, Candace Bramson, Lili Jiang, Feng Dai, Eva Szigethy, John Mellors, Barbara Methe, Frank Sciurba, Seyed Mehdi Nouraie, Alison Morris

## Abstract

**Objective:** To characterize subphenotypes of self-reported symptoms and outcomes(SRSOs) in Post-acute sequelae of COVID-19(PASC).

**Design:** Prospective, observational cohort study of PASC subjects.

**Setting:** Academic tertiary center from five clinical referral sources.

**Participants:** Adults with COVID-19 ≥ 20 days before enrollment and presence of any new self-reported symptoms following COVID-19.

**Exposures:** We collected data on clinical variables and SRSOs via structured telephone interviews and performed standardized assessments with validated clinical numerical scales to capture psychological symptoms, neurocognitive functioning, and cardiopulmonary function. We collected saliva and stool samples for quantification of SARS-CoV-2 RNA via qPCR.

**Primary and Secondary outcomes of measure:** Description of PASC SRSOs burden and duration, derivation of distinct PASC subphenotypes via latent class analysis (LCA), and relationship between viral load with SRSOs and PASC subphenotypes.

**Results:** Baseline data for 214 individuals were analyzed. The study visit took place at a median of 197.5 days after COVID-19 diagnosis, and participants reported ever having a median of 9/16 symptoms (interquartile range 6-11) after acute COVID, with muscle-aches, dyspnea, and headache being the most common. Fatigue, cognitive impairment, and dyspnea were experienced for a longer time. Participants had a lower burden of active symptoms (median 3, interquartile range 1-6) than those ever experienced (p<0.001). Unsupervised LCA of symptoms revealed three clinically-active PASC subphenotypes: a high burden constitutional symptoms (21.9%), a persistent loss/change of smell and taste (20.6%), and a minimal residual symptoms subphenotype (57.5%). Subphenotype assignments were strongly associated with self-assessments of global health, recovery and PASC impact on employment (p<0.001). Viral persistence (5.6% saliva and 1% stool samples positive) did not explain SRSOs or subphenotypes.

**Conclusions:** We identified distinct PASC subphenotypes and highlight that although most symptoms progressively resolve, specific PASC subpopulations are impacted by either high burden of constitutional symptoms or persistent olfactory/gustatory dysfunction, requiring prospective identification and targeted preventive or therapeutic interventions.

**Strengths and Limitations of this study:**

1. Prospective cohort study with inclusive patient population with PASC symptomatology from different clinical sources and index severity of COVID-19.
2. Structured telephone interviews and standardized assessments with validated clinical numerical scales
3. Unsupervised clustering analysis for data-driven derivation of PASC subphenotypes.
4. Analyses based on self-reported symptoms and outcomes, but not on physiologic or imaging measurements.
5. Non-invasive biospecimen for analysis of viral persistence may have missed viral signal in deep-seeded tissues.

**Key Points:** *Question:* Are there distinct subphenotypes of self-reported symptoms and outcomes in subjects with PASC and what if so, which factors predict them?

*Findings:* This prospective observational cohort study identified three distinct PASC clusters, comprising a high burden cluster with constitutional symptoms (21.9%), a cluster characterized by persistent loss/change of smell and taste (20.6%), and a minimal residual symptoms cluster (57.5%).

*Meaning:* PASC subphenotypes offer insights into the symptoms and outcomes experienced by individuals, and provide a framework for targeted study of preventive and therapeutic interventions.

## Introduction

The COVID-19 pandemic has had a profound global impact on public health. The multi-organ involvement of SARS-CoV-2 infection highlights the critical need to understand the post-acute sequelae of SARS-CoV-2 (PASC) and the long-term consequences on patients’ well-being and functionality.^1^ PASC, commonly referred to as long COVID, encompasses a wide range of clinical definitions, presentations and diverse trajectories.^2–4^ While most COVID-19 patients recover from their acute illness within a few weeks, PASC is estimated to affect 10-20% of COVID-19 survivors across all ages.^2,3,5,6^ Notably, PASC can manifest in patients with severe acute COVID-19 as well as those with milder initial disease, with up to 4 million Americans unable to return to work, regardless of the severity of their acute illness, according to many reports.^2,7^ Postulated mechanisms for PASC include viral, host, environmental and treatment factors.^8,9^ The absence of standardized phenotyping and systematic biological investigations has resulted in gaps in understanding prognosis for PASC and in developing effective preventive or therapeutic strategies.

We leveraged our clinical and research infrastructure to characterize subphenotypes of self-reported symptoms and outcomes (SRSOs) in subjects with PASC, identify factors associated with persistent PASC phenotypes, and investigate mechanisms of biological heterogeneity related to viral persistence.

## Methods

### Study Cohort

We conducted the Post-COVID Impairment Phenotyping and Outcomes [**Post-CIPO**] study, a prospective, observational cohort study with longitudinal follow-up of adult (≥18 years old) subjects with PASC-related SRSOs. The study was approved by the University of Pittsburgh IRB (STUDY21010001). We used an inclusive case definition of PASC, defined as the experience of any new or persistent symptoms for at least 20 days following a documented COVID-19 illness by positive qPCR. We enrolled patients from five different sources (see Supplement for details), classified as inpatients vs. outpatients at the time of COVID-19 illness. Following informed consent, we conducted a baseline study visit via structured telephone interviews during which we collected data on demographics, comorbid conditions, timeline of the previous COVID-19 illness(es), vaccinations and treatments received, and then types/duration/severity of 16 PASC-related symptoms. We selected the 16 specific symptoms based on expert input and knowledge on PASC symptomatology at the time of cohort inception. These symptoms included fever, chills, muscle aches, “runny nose”, sore throat, cough, dyspnea (“shortness of breath”), nausea or vomiting, headache, abdominal pain, diarrhea, loss/change of smell, loss/change of taste, cognitive impairment (“brain fog”), fatigue, and chest issues (pain or palpitations), with detailed questionnaires provided in the Appendix. We conducted standardized assessments with validated clinical numerical scales to capture the following domains of function and symptomatology: i) psychological symptoms: Generalized Anxiety Scale-7 [GAD7] for anxiety;^10^ patient health questionnaire-9 [PHQ9] for depression;^11^ insomnia severity index [ISI] for insomnia^12^, ii) neurocognitive functioning: Montreal Cognitive Assessment / MoCA-BLIND,^13^ and iii) cardiopulmonary function: Modified Medical Research Council [MMRC] Dyspnea scale.^14^ We also asked participants questions for self-assessment of their overall PASC outcomes in terms of global health, recovery, and impact on employment. For the purposes of this study, only baseline visit data were analyzed.

### Biospecimen acquisition and molecular analyses

Following the study visit, subjects self-collected stool and saliva samples at a single time point that were stored in nucleic acid preservation media and mailed to our laboratory. We aliquoted specimens and stored them at -80°C until conduct of experiments. We quantified SARS-CoV-2 viral load in available biospecimens with 1-step quantitative real-time-PCR of the SARS-CoV-2 N gene and human RNaseP gene.^15,16^

### Statistical analyses

To examine for presence of distinct subphenotypes (classes) of SRSOs, we conducted unsupervised classification with Latent Class Analysis (LCA) of self-reported symptoms at baseline visit in two separate analyses:^17^ i) we used symptoms experienced at any time post-COVID-19 (“ever-experienced”) as input variables to examine retrospectively for “epidemiologic clusters” (LCA-1), and ii) we used active symptoms at the time of the baseline visit to stratify subjects into “clinically active clusters” (LCA-2). We examined model fit performance by calculating membership probability, entropy, and the parametric bootstrapped log likelihood ratio between different classes, as well as a clinical relevance criterion of ensuring that each class has at least 5% of observations from the cohort. Results from the quantitative scales (GAD7, PHQ9, ISI, MoCA-BLIND, MMRC) were then mapped to each cluster across LCA-1 and LCA-2 analyses for a descriptive assessment of those data by cluster assignment. To examine whether cluster membership could be predicted by baseline clinical covariates, we used least absolute shrinkage and selection operator (LASSO) regression models (with 10 folds validation) with clinical covariates as predictors and clusters as outcomes. We conducted subgroup analyses for inpatients and outpatients, separately. We performed non-parametric comparisons for continuous (described as median and interquartile range – IQR) and categorical variables between different groups and reported the nominal p-values for all tests performed, without adjustments for multiple comparisons. From available stool and saliva samples, we examined differential levels of SARS-CoV-2 RNA in stool and saliva samples by SRSOs and PASC subphenotypes. We conducted analyses in R (v4.2.0), STATA 17.0 and Mplus 8.8.

## Results

### Cohort Description

From March 2021 through January 2023, we enrolled a feasibility cohort 214 individuals with PASC through various referral sources, when subjects reported symptoms consistent with PASC or sought care for PASC (Table 1). The study visit took place at a median of 197.5 (IQR 143.0-323.2) days following COVID-19 diagnosis. Patients who were hospitalized during their acute COVID-19 illness (32% inpatients, Table 1) were older, with higher burden of comorbid conditions, and were interviewed closer to their COVID-19 diagnosis compared to outpatients (all p<0.01, Table 1). The UPMC Post-COVID clinic was the most common referral source to the Post-CIPO study (31.8%, Table S1). Participants enrolled through the Post-COVID clinic or who were self-referred to the study were younger and with fewer comorbid conditions compared to patients referred from inpatient or outpatient studies of acute COVID-19 or referred by physicians for PASC (Table S1).

**Table 1:** Clinical characteristics of the 214 subjects included with Post Acute Sequelae (PASC) of COVID-19, stratified by inpatient vs. outpatient status during acute COVID-19. We present continuous variables as median and interquartile range (IQR) and categorical variables as number (%). We compared continuous variables with Wilcoxon tests and categorical variables with Fisher’s exact tests. We consider p<0.05 as statistically significant.

| Variable | All | Outpatients | Inpatients | P Value |
| --- | --- | --- | --- | --- |
| **Participants** | 214 | 143 | 71 |  |
| **Age (median, [IQR]), years** | 49.9 [38.5, 62.3] | 44.1 [34.4, 54.6] | 62.0 [51.6, 68.5] | <0.01 |
| **Men (%)** | 57 (26.6) | 30 (21.0) | 27 (38.0) | 0.01 |
| **Whites (%)** | 196 (91.6) | 132 (92.3) | 64 (90.1) | 0.78 |
| **Body Mass Index (BMI) (median [IQR])** | 30.0 [25.1, 34.7] | 28.9 [24.1, 33.6] | 31.3 [27.0, 36.6] | 0.01 |
| **Inpatients (%)** | 71 (33.2) | 0 (0.0) | 71 (100.0) | <0.01 |
| **No college-level degree (%)** | 101 (47.2) | 56 (39.2) | 45 (63.4) | <0.01 |
| **Hypertension (%)** | 75 (35.0) | 43 (30.1) | 32 (45.1) | 0.04 |
| **Diabetes (%)** | 32 (15.0) | 8 (5.6) | 24 (33.8) | <0.01 |
| **Coronary artery disease (%)** | 8 (3.7) | 3 (2.1) | 5 (7.0) | 0.16 |
| **Congestive Heart Failure (%)** | 7 (3.3) | 0 (0.0) | 7 (9.9) | <0.01 |
| **Stroke (%)** | 9 (4.2) | 1 (0.7) | 8 (11.3) | <0.01 |
| **Atrial fibrillation (%)** | 13 (6.1) | 3 (2.1) | 10 (14.1) | <0.01 |
| **Obstructive sleep apnea (%)** | 54 (25.2) | 23 (16.1) | 31 (43.7) | <0.01 |
| **Obstructive airways disease\* (%)** | 60 (28.0) | 35 (24.5) | 25 (35.2) | 0.14 |
| **History of cancer (%)** | 11 (5.1) | 6 (4.2) | 5 (7.0) | 0.58 |
| **History of immunosuppression (%)** | 33 (15.4) | 15 (10.5) | 18 (25.4) | 0.01 |
| **Anemia (%)** | 35 (16.4) | 16 (11.2) | 19 (26.8) | 0.01 |
| **Ever smoker (%)** | 80 (37.4) | 47 (32.9) | 33 (46.5) | 0.07 |
| **Vaccinated for Influenza (%)** | 140 (65.4) | 97 (67.8) | 43 (60.6) | 0.37 |
| **Vaccinated for COVID-19 (%)** | 122 (57.0) | 81 (56.6) | 41 (57.7) | 0.99 |
| **No of COVID-19 vaccinations (median [IQR])** | 2.0 [0.0, 3.0] | 2.0 [0.0, 3.0] | 2.0 [0.0, 3.0] | 0.44 |
| **Antiviral treatment during acute COVID-19 (%)** | 33 (15.4) | 4 (2.8) | 29 (40.8) | <0.01 |
| **Prevalent SARS-CoV-2 variant during each acute infection period** |  |  |  | <0.01 |
| **Wild Type** | 73 (34.1) | 32 (22.4) | 41 (57.7) |  |
| **Alpha** | 73 (34.1) | 53 (37.1) | 20 (28.2) |  |
| **Delta** | 37 (17.3) | 29 (20.3) | 8 (11.3) |  |
| **Omicron** | 31 (14.5) | 29 (20.3) | 2 (2.8) |  |
| **Days since acute COVID-19 infection date (median [IQR])** | 197.5 [143.0, 323.2] | 221.0 [155.5, 350.5] | 161.0 [123.5, 238.0] | <0.01 |

### Baseline SRSOs

We first examined symptoms “ever-experienced” post-COVID-19 and how long they lasted. Overall, participants endorsed a median of nine symptoms (IQR 6-11) out of the total 16 symptoms in our questionnaire, with muscle aches, dyspnea and headache being the most common ones (reported by >70% of participants). Distributions of the duration of these symptoms (expressed as estimated number of days for each symptom post-COVID) were highly right skewed (Figure 1A), with fatigue, cognitive impairment and dyspnea experienced for longer periods of time among those who reported these symptoms. At the time of baseline visit, participants endorsed a lower burden of active symptoms (median 3 [1-6]) compared to those “ever-experienced” symptoms (median 9 [6-11], paired Wilcoxon test p<0.001, Figure 1B). The most common active symptoms at time of baseline visit were dyspnea, fatigue and cognitive impairment (reported by >40% of participants, Figure 1C). The numerical scales examined from baseline visit (Figure 1D) showed that more than half of participants reported mild anxiety or depression (GAD7>4 or PHQ9>4, respectively) while 42.5% reported moderate or severe insomnia and 43.6% reported clinically significant dyspnea (MMRC≥2). For objective neurocognitive testing, 34.4% had an abnormal MoCA-BLIND test (<18). For the self-assessment of outcomes, 39.7% deemed their general health as excellent or very good, 20% felt that they had fully recovered from COVID-19, whereas 49.6% reported that their employment ability had been affected to various degrees by PASC (Figure 1E). We found significant associations of clinical covariates with active symptoms and clinical scales (Figures S1-6), with COVID-19 vaccination associated with lower scores for depression, anxiety and insomnia, and higher odds for feelings of full recovery (Figure S7).

**Figure 1:**
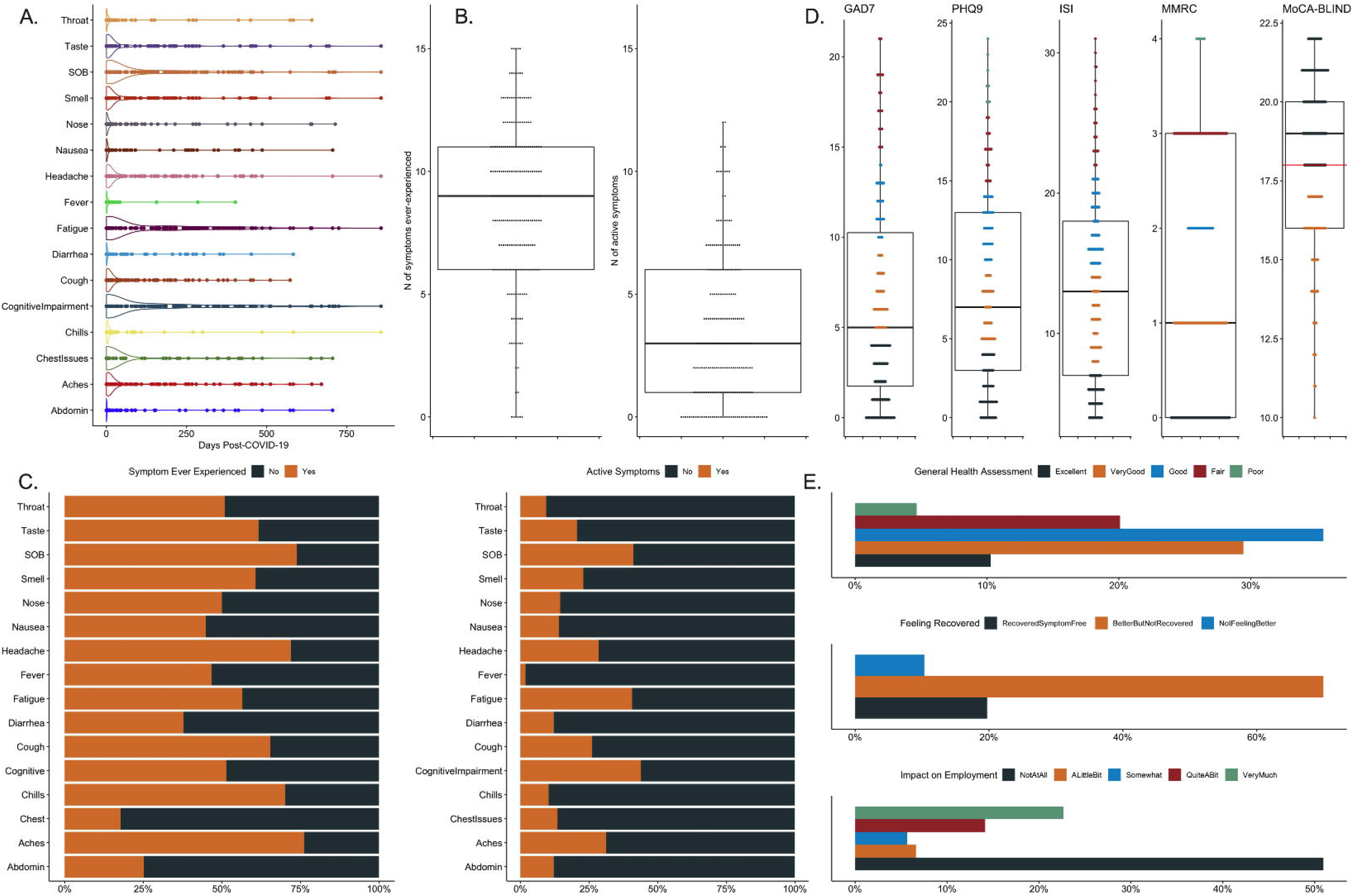
Self-reported symptoms and outcomes among 214 subjects with Post-Acute Sequelae of COVID-19 (PASC) at baseline visit. A. Distribution of the duration (in days) of each of the 16 interviewed symptoms post COVID-19. B. Subjects reported a median of 9 (interquartile range [IQR] 6-11) “ever-experienced” symptoms and a median of 3 (IQR 1-6) active symptoms at baseline visit. C. Stacked bar showing the proportions of presence (“Yes” in orange) vs. absence (“No” in gray) for each of the 16 interviewed symptoms, with “ever-experienced” symptoms shown in the left panel and active symptoms in the right panel. D. Distributions of the numerical scales examined in the baseline visit: Generalized Anxiety Scale-7 (GAD7) for anxiety; patient health questionnaire-9 (PHQ9) for depression; insomnia severity index (ISI) for insomnia, Montreal Cognitive Assessment / MoCA-BLIND for neurocognitive functioning and Modified Medical Research Council (MMRC) Dyspnea scale for cardiopulmonary function. An abnormal MoCA-BLIND test was defined as score of less than 18 (red line). E. Self-assessed outcomes of general health (top panel), recovery from COVID-19 (middle panel) and impact of COVID-19 on employment (bottom panel).

### PASC subphenotypes

In the “epidemiologic cluster” analysis, we conducted LCA by using symptoms “ever-experienced” post-COVID as input variables (LCA-1). LCA-1 revealed that a 3-class model offered optimal fit, with about equal distribution between the three classes (clusters, Table S2). Cluster 1 had markedly higher symptom burden, followed by cluster 2, whereas cluster 3 was an overall low symptom burden PASC subgroup (Figure S8A). More than 50% of cluster 1 subjects had experienced 15/16 symptoms (except for chest issues), whereas nearly all cluster 2 subjects reported loss/change of smell and taste (Figure S8B). Cluster 1 subjects had higher scores in scales for anxiety, depression, and insomnia but no difference in neurocognitive scale (Figure S8C). We found no significant differences between clusters by baseline covariate (Table S2), but cluster membership was significantly associated with the self-assessed impact of PASC on employment (Figure S9).

Next, we performed a “clinically active” LCA by using active symptoms (LCA-2). LCA-2 also revealed that a 3-class model offered optimal fit, but subjects were now less evenly distributed. Cluster 1 contained patients with higher symptom burden than cluster 2, but cluster 2 was distinguished by its high proportion of patients reporting a persistent loss of taste and smell (Figure 2A). Cluster 1 subjects had higher scores for anxiety, depression, and insomnia scales, but no difference in the neurocognitive MoCA-BLIND scale (Figure 2C), despite 81% self-reporting cognitive impairment. Notwithstanding the lower burden of active symptoms in cluster 2 compared to cluster 1 (Figure 2A, p<0.001), clusters 1 and 2 had similarly poor outcomes in terms of subjective recovery and impact on employment (Figure S10).

**Figure 2:**
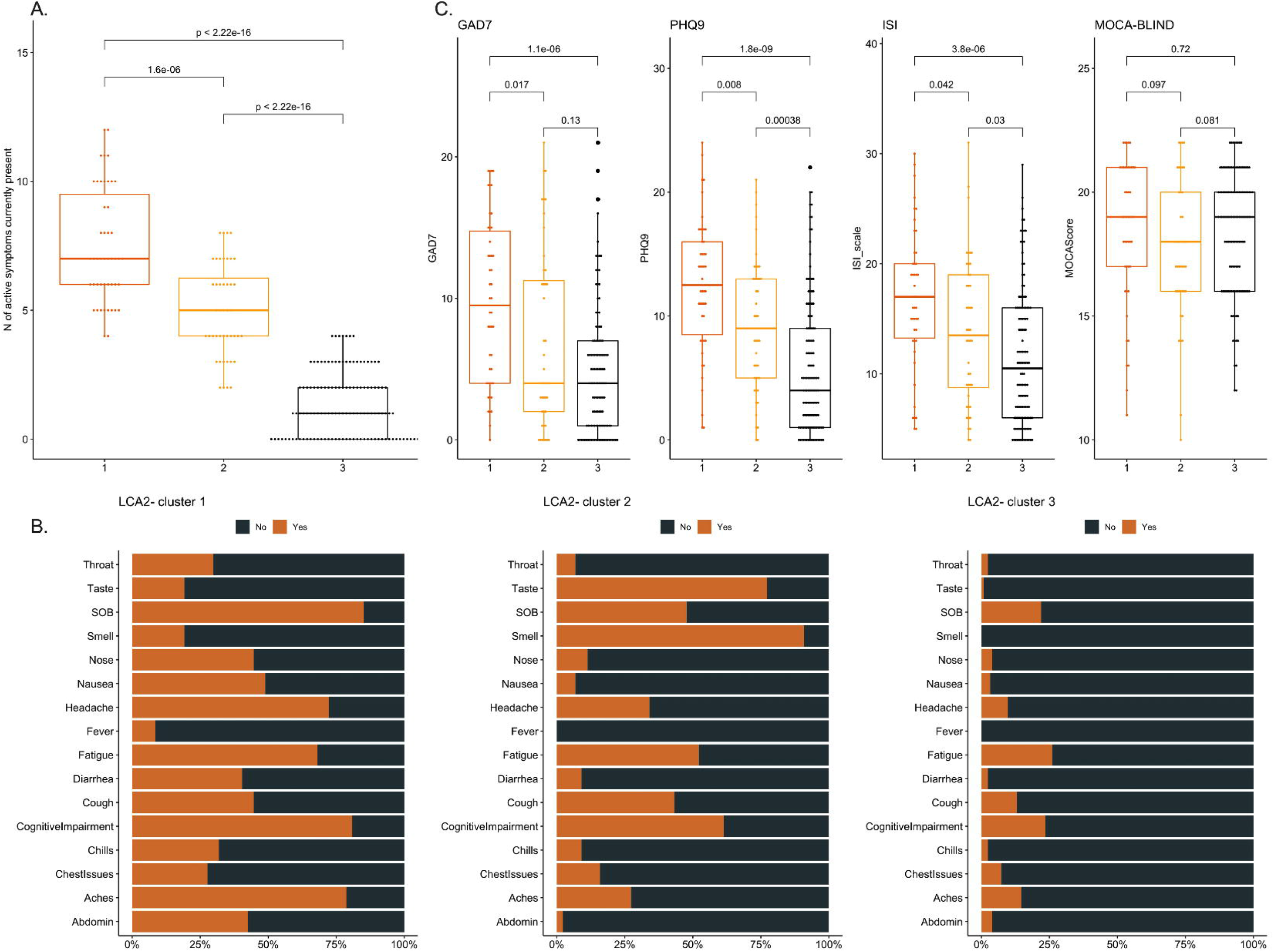
Distributions of symptoms and numerical scales by the “clinically-active” subphenotypes of Post-Acute Sequelae of COVID-19 (PASC). We conducted latent class analysis (LCA) by using the 16 active symptoms at baseline visit as input variables (“LCA2”). A. Cluster 1 subjects had significantly higher number of active symptoms compared to cluster 2 subjects, who in turn had much higher number of symptoms compared to cluster 3. B. Stacked bar showing the proportions of presence (“Yes” in orange) vs. absence (“No” in gray) for each of the 16 interviewed symptoms for each of the three clusters. C. Cluster 1 subjects had much higher scores for the numerical scales Generalized Anxiety Scale-7 (GAD7) for anxiety, patient health questionnaire-9 (PHQ9) for depression and insomnia severity index (ISI) for insomnia, but no difference in the Montreal Cognitive Assessment / MoCA-BLIND score for neurocognitive functioning.

We then integrated the results from the two clustering approaches (LCA-1 and LCA-2) to understand how subjects transitioned over time from the epidemiologic (LCA-1) to the “clinically-active” (LCA-2) classifications. LCA-1 and LCA-2 cluster memberships were strongly associated (McNemar’s paired test p<0.001), with notable transitions between clusters (Figure 3A). A majority of cluster 3 subjects in the LCA-1 (88.9%) were also classified as cluster 3 by LCA-2, representing a population with consistently low number of PASC symptoms (“Low Burden PASC”). Among subjects placed in clusters 1 and 2 by LCA-1, we noted that 41.8% of the multi-symptomatic cluster 1 and 46.4% of the predominantly impaired taste/smell cluster 2 were classified as cluster 3 by LCA-2, representing a population who was highly symptomatic at some point post-COVID, but reported a low symptom burden by the time of the baseline visit. Thus, we considered such subjects transitioning to LCA-2 cluster 3 as representative of “Resolved PASC”. Subjects classified as cluster 1 and 2 by LCA-2 represented a population of “PASC Persisters” (42.5%), either for the continued impairment of taste/smell in cluster 2 or for multiple symptoms in cluster 1 (Figure 3A, Table S4). “PASC Persisters” had significantly worse clinical scales for anxiety, depression and insomnia than “Low Burden” or “PASC Resolvers”, but no difference in the neurocognitive MoCA-BLIND scale (Figure 3B). “PASC Persisters” reported significantly worse outcomes for subjective recovery and impact on employment compared to “Low Burden” or “PASC Resolvers” (p<0.001, Figure 3C). We found no significant differences in baseline covariates between these integrative PASC clusters, other than “PASC Persisters” having a lower proportion of vaccination (49.5%) compared to “Low Burden” or “PASC Resolvers” (66.1% and 59.7%, respectively; p<0.001, Table 2).

**Figure 3:**
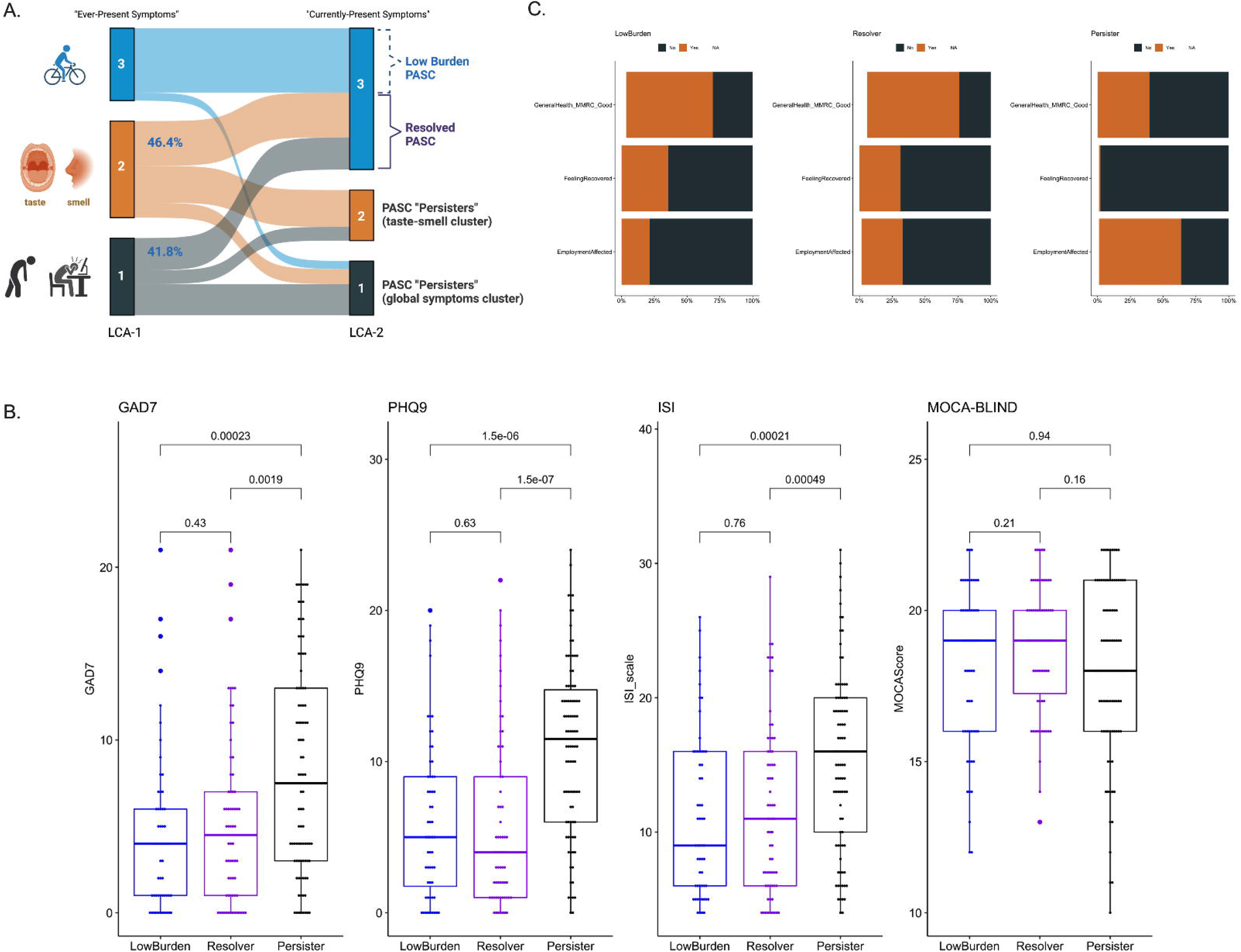
Integrative clustering analysis revealed a subset of PASC “Persisters” with high symptom burden, poor self-assessment outcomes and high numerical scales for anxiety, depression and insomnia. A. Transitions between the two clustering approaches (LCA-1 and LCA-2). LCA-1 analyzed “ever-experienced” symptoms (epidemiologic subphenotyping) whereas LCA-2 analyzed active symptoms (“clinically-active” subphenotyping). Most cluster 3 subjects by LCA-1 (88.9%) were also classified as cluster 3 by LCA-2 (“Low Burden PASC”). Among subjects in clusters 1 and 2 by LCA-1, 41.8% of the multi-symptomatic cluster 1 and 46.4% of the taste/smell predominant cluster 2 were classified as cluster 3 by LCA-2 (“Resolved PASC”). Subjects classified as cluster 1 and 2 by LCA-2 represented a population of “PASC Persisters”, either for taste/smell predominant cluster 2 or the multi-symptomatic cluster 1. B. “PASC” Persisters” had significantly worse scales for anxiety, depression and insomnia than “Low Burden” or “PASC Resolvers”, but no difference in the neurocognitive MoCA-BLIND scale. C. “PASC Persisters” reported significantly worse outcomes for global health assessment, subjective recovery and impact on employment compared to “Low Burden” or “PASC Resolvers” (p<0.001).

**Table 2:** Clinical characteristics by integrative clusters of methods LCA-1 and LCA-2 through which we defined the subphenotypes of Persisters, Resolvers or Low Burden PASC subjects.

| Variable | Low Burden | Resolver | Persister | P Value |
| --- | --- | --- | --- | --- |
| **Participants** | 56 | 67 | 91 |  |
| **Age (median [IQR])** | 56.6 [36.3, 67.8] | 49.4 [33.9, 57.9] | 50.0 [41.4, 61.7] | 0.08 |
| **Men (%)** | 18 (32.1) | 19 (28.4) | 20 (22.0) | 0.37 |
| **Whites (%)** | 48 (85.7) | 61 (91.0) | 87 (95.6) | 0.11 |
| **Body Mass Index (BMI) (median [IQR])** | 30.0 [25.6, 34.4] | 29.3 [25.3, 34.1] | 30.2 [25.1, 35.3] | 0.77 |
| **Inpatients (%)** | 27 (48.2) | 20 (29.9) | 24 (26.4) | 0.02 |
| **No college-level degree (%)** | 28 (50.0) | 26 (38.8) | 47 (51.6) | 0.25 |
| **Hypertension (%)** | 23 (41.1) | 20 (29.9) | 32 (35.2) | 0.43 |
| **Diabetes (%)** | 11 (19.6) | 7 (10.4) | 14 (15.4) | 0.36 |
| **Coronary Artery Disease (%)** | 3 (5.4) | 1 (1.5) | 4 (4.4) | 0.48 |
| **Congestive Heart Failure (%)** | 3 (5.4) | 2 (3.0) | 2 (2.2) | 0.57 |
| **Stroke (%)** | 3 (5.4) | 2 (3.0) | 4 (4.4) | 0.8 |
| **Atrial Fibrillation (%)** | 5 (8.9) | 3 (4.5) | 5 (5.5) | 0.56 |
| **OSA (%)** | 13 (23.2) | 15 (22.4) | 26 (28.6) | 0.62 |
| **Obstructive airways disease (%)** | 11 (19.6) | 19 (28.4) | 30 (33.0) | 0.22 |
| **History of Cancer (%)** | 3 (5.4) | 3 (4.5) | 5 (5.5) | 0.96 |
| **History of Immunosuppression (%)** | 9 (16.1) | 8 (11.9) | 16 (17.6) | 0.62 |
| **Anemia (%)** | 9 (16.1) | 11 (16.4) | 15 (16.5) | 1 |
| **Ever smoker (%)** | 23 (41.1) | 19 (28.4) | 38 (41.8) | 0.18 |
| **Vaccinated for Influenza (%)** | 37 (66.1) | 46 (68.7) | 57 (62.6) | 0.73 |
| **Vaccinated for COVID-19 (%)** | 37 (66.1) | 40 (59.7) | 45 (49.5) | 0.12 |
| **No of COVID-19 vaccinations, (median [IQR])** | 2.0 [0.0, 3.0] | 2.0 [0.0, 3.0] | 0.0 [0.0, 3.0] | 0.02 |
| **Antiviral Treatment during acute COVID-19 (%)** | 12 (21.4) | 11 (16.4) | 10 (11.0) | 0.23 |
| **Prevalent SARS-CoV-2 variant during each acute infection period** |  |  |  | <0.01 |
| **Wild Type** | 13 (23.2) | 28 (41.8) | 32 (35.2) |  |
| **Alpha** | 4 (7.1) | 13 (19.4) | 20 (22.0) |  |
| **Delta** | 10 (17.9) | 13 (19.4) | 8 (8.8) |  |
| **Omicron** | 29 (51.8) | 13 (19.4) | 31 (34.1) |  |
| **Days since acute COVID-19 infection date, (median [IQR])** | 186.5 [140.8, 279.0] | 181.0 [125.5, 319.0] | 214.0 [146.5, 372.0] | 0.24 |

### Clinical predictors of PASC subphenotypes

Following derivation of PASC subphenotypes, we examined whether clinical covariates available at the time of COVID-19 diagnosis may predict PASC cluster membership with LASSO models. LASSO regression models found no significant predictors for the epidemiologic clusters (LCA-1). For the clinically active clusters (LCA-2), LASSO regression revealed that higher BMI, lower education level, history of anemia and autoimmune disease increased the risk for the multi-symptomatic cluster 1 membership, whereas history of COVID-19 hospitalization, COVID vaccination and infection with the delta variant were protective against cluster 1 membership (Table S4).

### Viral persistence and SRSOs

We measured SARS-CoV-2 RNA (vRNA) load in 103 saliva samples and 101 stool samples available. We found detectable vRNA in 6 saliva (5.8%) and 1 (1.0%) stool sample. Therefore, we were able to perform exploratory analyses comparing viral positive and negative samples for saliva only (6 vs. 97 samples, respectively, Table S5). Subjects with viral positive saliva samples had donated samples closer to their acute COVID-19 diagnosis (median of 133.0 vs. 188.0 days, p<0.001), were more likely to have received antiviral treatment during acute COVID-19 (p=0.04), but otherwise had no significant differences in individual symptoms or cluster membership compared to subjects with negative viral load (Table S6).

## Discussion

Our study examined subphenotypes of SRSOs in an inclusive cohort of PASC subjects. Our findings demonstrate the heterogeneous nature of PASC, with varying symptomatology, functional impairments, and time course. Through unsupervised analysis of 16 self-reported symptoms, we identified three distinct PASC subphenotypes. Our results suggest that individuals evaluated for PASC at different time intervals after COVID-19 comprise discrete subpopulations of symptom burden and duration, with likely differing underlying disease mechanisms and care needs. The first subphenotype was characterized by high symptom burden (cluster 1), whereas the second subphenotype was distinguished by disturbances of smell and taste (cluster 2). Most subjects showed low symptom burden or had resolved PASC by the time of baseline visit (cluster 3). Most subjects in clusters 1 and 2 felt that they have not recovered from PASC, which impacted their employment. We investigated oral and gastrointestinal viral persistence as potential biological mechanisms for PASC, but overall found a low prevalence of detectable viral RNA in saliva and stool samples.

To better understand how COVID-19 survivors were feeling and functioning, we conducted structured telephone interviews enrolling individuals at various time intervals post COVID-19. PASC subjects reported a median of nine symptoms that were “ever experienced” since contracting COVID-19, with individual symptom durations being notably long. By the time of the baseline visit, which was conducted at a median of 197.5 days post-COVID, most symptoms had resolved, leading to a phenotype of low symptom burden or resolved PASC for most subjects. Nevertheless, 42.5% of participants displayed persistent PASC (clusters 1 and 2 in LCA-2), with poor self-assessment of health and recovery, as well as adverse impact on employment. PASC “Persisters” had higher scores for anxiety, depression and insomnia, and reported much higher prevalence of cognitive impairment, despite showing similar scores on objective neurocognitive scale testing, which may have not been detailed enough to capture more subtle neurocognitive deficits. Therefore, even among patients who are referred or personally seek care for PASC at some point after COVID-19, it is the subset of “Persisters” (42.5% in our cohort) that has high symptom burden and impaired functional outcomes that may warrant specific evaluation and treatment.

Among PASC “Persisters”, we identified a distinct cluster with disturbances of smell and taste. Such disturbances varied from decreased or absent smell or taste to distorted or putrid sensations, which are common during acute COVID-19, but are also increasingly recognized as components of PASC with variable recovery trajectories seen in different studies.^4,18^ A recent large multicenter cohort study identified four PASC clusters, of which one cluster was defined by the persistence of smell and taste disturbances in all included subjects,^3^ highlighting the external validity of our clustering analysis. The proposed mechanisms of persistent olfactory and/or gustatory dysfunction involve conductive and sensorineural deficits, likely due to acute mucosal or neuronal damage during acute COVID-19.^19^ Therefore, these patients represent a distinct PASC subtype, which will require much different biological study and testing of interventions compared to other PASC clusters with constitutional symptoms and cognitive impairments.^3^

To improve subject identification for PASC studies enriched for highly symptomatic subjects, we conducted LASSO modeling to define predictors of PASC subphenotypes. We selected clinical variables that were easily accessible from EMR reviews or subject interviews, as well as variables that had face value for PASC associations based on univariate analyses with SRSOs or LCA clusters. We found that a simple clinical model based on BMI, education level, anemia, immune disease, hospitalization, vaccination and delta variant infection accurately predicted cluster 1 membership by LCA-2. Concordant to recent evidence,^3,20^ our analyses also showed that vaccination was associated with lower probability of assignment to a multi-symptomatic PASC cluster. Although external validation in larger datasets is required to establish generalizability of the prognostic value of such models, our analysis suggests that clinical variables available at the time of acute COVID-19 infection may aid in prioritizing follow-up and study enrollment for subjects who are more likely to develop highly symptomatic and persistent PASC.

Our molecular analyses of non-invasive biospecimens with a sensitive qPCR assay for SARS-CoV-2 RNA ^15,16^ showed overall low prevalence of viral persistence (5.8% for saliva and 1% for stool samples). The six subjects with viral positive saliva samples had donated samples at a median of 133.0 days following their acute COVID-19 diagnosis, and none of them reported a COVID-19 re-infection. Although such viral RNA detection does not prove ongoing viral replication or infectivity, we note that the studies of inpatients with COVID-19 have shown much shorter durations of saliva viral positivity (e.g. a median of 18.0 days).^21^ The small sample size of positive saliva samples in our cohort did not allow us to perform robust testing of associations between viral load and PASC SRSOs or subphenotypes. We could not draw any inferences about viral persistence in other body reservoirs. However, our results allow us to conclude that viral RNA quantification in non-invasive biospecimens is unlikely to explain the observed PASC heterogeneity, and that future study of PASC needs to consider different biospecimens for viral persistence as well as additional biological mechanisms.^8^

Our study has several limitations. First, we relied solely on SRSOs and did not conduct objective physiologic tests, such as pulmonary function tests or neuro-imaging, which could have provided a more comprehensive understanding of PASC. Nonetheless, our analysis of self-reported symptom onset, duration, and severity provided important insights into the broad spectrum of symptomatology experienced post COVID-19. Second, our study included participants from five different sources, which may have introduced selection biases and affected the generalizability of our findings. For instance, we observed lower PASC symptom burden in hospitalized patients who had more severe acute COVID-19 than outpatients. However, this does not suggest that severe acute COVID-19 is protective against PASC, but rather that the outpatients enrolled in our PASC cohort are likely to have a higher burden of PASC symptoms. Therefore, we urge caution when interpreting the observed associations. Finally, it is important to recognize that the PASC burden and impact on individuals’ lives can vary widely, and our findings may not be generalizable to all PASC patients.

In summary, our study sheds light on the clinical heterogeneity of PASC. We identified distinct PASC subphenotypes driven by symptomatology type and burden. We show that viral persistence in non-invasive biospecimens has low prevalence and does not explain SRSOs or PASC subphenotypes. Our approach to PASC subphenotyping provides a reproducible framework for capturing a wide spectrum of SRSOs and identifying clinical subtypes with adverse impact on patient-centered endpoints. Future research on PASC should focus on developing and validating predictive models for timely identification of COVID-19 survivors who are at high risk of persistent PASC, as well as targeting mechanistic study and interventional trials in distinct subsets of patients with either high burden of constitutional symptoms or persistent olfactory/gustatory dysfunction.

## Ethics approval and consent to participate

The University of Pittsburgh Institutional Review Board (IRB) approved the study protocol STUDY21010001. We obtained written or electronic informed consent by all participants in accordance with the Declaration of Helsinki.

## Consent for publication

We obtained the necessary participant consent and the appropriate institutional forms have been archived. Any patient/participant/sample identifiers included were not known to anyone outside the research group so cannot be used to identify individuals.

## Availability of data and material

All data generated or analyzed during this study are included in this article and its supplementary information files.

## Author Contributions

i. Conception or design of the work; or the acquisition, analysis, or interpretation of data for the work; AND
ii. Drafting the work or revising it critically for important intellectual content; AND
iii. Final approval of the version to be published; AND
iv. Agreement to be accountable for all aspects of the work in ensuring that questions related to the accuracy or integrity of any part of the work are appropriately investigated and resolved.

Georgios D. Kitsios: I, II, III, IV

Shawna Blacka: The acquisition, analysis, or interpretation of data for the work; II, III, IV

Jana Jacobs: The acquisition, analysis, or interpretation of data for the work; II, III, IV

Taaha Mirza: The acquisition, analysis, or interpretation of data for the work; II, III, IV

Asma Naqvi: The acquisition, analysis, or interpretation of data for the work; II, III, IV

Heather Gentry: The acquisition, analysis, or interpretation of data for the work; II, III, IV

Cathy Kessinger: The acquisition, analysis, or interpretation of data for the work; II, III, IV

Xiaohong Wang: The acquisition, analysis, or interpretation of data for the work; II, III, IV

Konstantin Golubykh: The acquisition, analysis, or interpretation of data for the work; II, III, IV

Hafiz Muhammad Siddique Qurashi: The acquisition, analysis, or interpretation of data for the work; II, III, IV

Akash Dodia: The acquisition, analysis, or interpretation of data for the work; II, III, IV

Michael Risbano: The acquisition, analysis, or interpretation of data for the work; II, III, IV

Michael Benigno: The acquisition, analysis, or interpretation of data for the work; II, III, IV

Birol Emir: The acquisition, analysis, or interpretation of data for the work; II, III, IV

Edward Weinstein: The acquisition, analysis, or interpretation of data for the work; II, III, IV

Candace Bramson: The acquisition, analysis, or interpretation of data for the work; II, III, IV

Lili Jiang: The acquisition, analysis, or interpretation of data for the work; II, III, IV

Feng Dai: The acquisition, analysis, or interpretation of data for the work; II, III, IV

Eva Szigethy: The acquisition, analysis, or interpretation of data for the work; II, III, IV

John Mellors: The acquisition, analysis, or interpretation of data for the work; II, III, IV

Barbara Methe: The acquisition, analysis, or interpretation of data for the work; II, III, IV

Frank Sciurba: The acquisition, analysis, or interpretation of data for the work; II, III, IV

Seyed Mehdi Nouraie: I, II, III, IV

Alison Morris: I, II, III, IV

## Data Availability

Availability of data and material: All data generated or analyzed during this study are included in this article and its supplementary information files.

