## Supplement for "Subphenotypes of Self-Reported Symptoms and Outcomes in Long COVID: a prospective cohort study with latent class analysis"

### **Study design**

We designed and conducted the Post-COVID Impairment Phenotyping and Outcomes [**Post-CIPO**] study, a prospective, observational cohort study with longitudinal follow-up of subjects with self-reported symptoms of “Long COVID” or Post-acute sequelae of SARS-CoV-2 infection (PASC). The study was approved by the University of Pittsburgh IRB (STUDY21010001). We included adults (≥18 years old) with documented prior COVID-19 illness at least 20 days prior to enrollment and presence of any new self-reported symptoms following COVID-19. We enrolled patients from five different sources (see Flowchart of Figure 1 for details):

a. Referrals from a post-COVID19 recovery clinic at the University of Pittsburgh Medical Center (UPMC) for patients examined for any persistent symptoms following COVID-19 (“Long COVID” clinic).
b. Direct physician referrals for patients with any persistent symptoms following COVID-19.
c. Follow-up of patients previously enrolled in an inpatient registry of COVID-19 (COVID INpatient Cohort [COVID-INC], STUDY20040036) following ≥ 20 days from hospital discharge. Details of the study procedures during the acute hospitalization of the COVID-INC cohort were previously described.[^2^](https://sciwheel.com/work/citation?ids=13814041&pre=&suf=&sa=0&dbf=0)
d. Follow-up of patients previously enrolled in an outpatient registry of COVID-19 (COVID-19 Outpatient Biobank [COB], STUDY20100419) following ≥ 20 days from enrollment to the COB study.
e. Patient self-referrals for enrollment through the Pitt+Me study at the University of Pittsburgh.

Based on our referral mechanism from different sources, we enrolled patients who at the time of the index COVID-19 had severe presentations to require hospitalization (referred to as *inpatients* at time of index COVID-19 illness) vs. those who had milder disease severity and experienced the COVID-19 index illness as *outpatients*. The terms *inpatients* vs. *outpatients* are therefore used to distinguish severity of the index COVID-19 illness (and not the status of patients at the timing of their enrollment to the **Post-CIPO** study).

#### Study Procedures:

a. Informed consent discussion and obtaining informed consent
b. Structured telephone interviews
c. Data extraction from electronic medical record
d. Biospecimen acquisition.

Following informed consent (a), we conducted a **baseline study visit** via structured telephone interviews (b) during which we collected the following data:

- demographic information,
- comorbid conditions,
- self-assessments of health status and functionality,
- smoking history exposure,
- timeline of the previous COVID-19 illness(es),
- vaccinations and treatments received,
- types/duration/severity of symptoms during the index COVID-19 illness and persistence of such symptoms at the time of the interview (detailed questionnaires provided in the Appendix).

We then conducted standardized assessments with validated clinical numerical scales to capture the following domains of functioning and symptomatology:

- psychological symptoms:
  - Generalized Anxiety Scale-7 [GAD7] for anxiety;[^3^](https://sciwheel.com/work/citation?ids=771330&pre=&suf=&sa=0&dbf=0)
  - Patient Health Questionnaire-9 [PHQ9] for depression;[^4^](https://sciwheel.com/work/citation?ids=1242607&pre=&suf=&sa=0&dbf=0)
  - Insomnia Severity Index [ISI] for insomnia[^5^](https://sciwheel.com/work/citation?ids=2306269&pre=&suf=&sa=0&dbf=0),
- neurocognitive functioning: Montreal Cognitive Assessment / MoCA-BLIND,[^6^](https://sciwheel.com/work/citation?ids=8629636&pre=&suf=&sa=0&dbf=0)
- cardiopulmonary function: Modified Medical Research Council [MMRC] Dyspnea scale.[^7^](https://sciwheel.com/work/citation?ids=1791826&pre=&suf=&sa=0&dbf=0)

All subjects completed a baseline visit at enrollment, and then were contacted for follow-up study visits at 1, 3-, 6-, 9- and 12-months post-enrollment. At follow-up visits, we collected updated information on any interval SARS-CoV-2 re-infection, update vaccination/treatments, and recorded symptoms at the time of the follow-up visits and clinical scale assessments (MoCA-BLIND; MMRC).

c. Data extraction from electronic medical record. We supplemented patient-reported information on vaccination and COVID-19 treatment administration by targeted review and data extraction for such variables from the electronic medical record at UPMC. Specifically for subjects enrolled from the inpatient COVID-INC study, we also extracted and recorded extensive clinical variables during the index hospitalization of COVID-19, including severity of illness by the World Health Organization (WHO) 10-point ordinal scale on presentation and on discharge, COVID-19 specific therapeutics (including corticosteroids and additional immunomodulators), respiratory support modalities, and clinical endpoints of 60-day survival and discharge destination for survivors. We also retrieved baseline chest radiography images at the time of hospital admission and quantified radiographic severity with the Radiographic Assessment Lung Edema (RALE) score, as previously described.[^2^](https://sciwheel.com/work/citation?ids=13814041&pre=&suf=&sa=0&dbf=0)

d. Biospecimen acquisition:

Following their baseline visit, subjects were mailed to their home address a kit for collection of stool and saliva samples, with provision of clear instructions.

- **Stool specimens** were self-collected using the DNA/RNA Shield Fecal Collection tubes (Zymo) for nucleic acid preservation and short-term (two to four weeks) storage at ambient temperature.
- **Saliva specimens** were self-collected using the OMNIgene·ORAL OM-505 devices. 2 mL of saliva were collected for nucleic acid preservation and short-term storage at ambient temperature.

Specimens were mailed to the University of Pittsburgh Center for Medicine and the Microbiome. Upon receipt, specimens were sub-aliquoted prior to long-term storage at -80°C.

#### Molecular Analyses:

To quantify the SARS-CoV-2 viral load in available biospecimens, we extracted total RNA from 0.35 mL of inactivated saliva and stool samples (when available) using the MagMax Viral Pathogens Kit (Thermofisher), and performed 1-step quantitative RT-PCR of the SARS-CoV-2 N gene and human RNaseP gene, as previously described, using the following N-specific primers (forward: 5′ -GTTTGGTG GACCCTCAGATT-3′ , reverse: 5′ -CGCAGTATTATTGGGT AAACCTTG-3′ , Probe: 5′ 6-FAM-TAACCAGAATGGAGAAC GCAGTGGG-3′ BHQ1).[^9,10^](https://sciwheel.com/work/citation?ids=12208453,13061298&pre=&pre=&suf=&suf=&sa=0,0&dbf=0&dbf=0)

***Quantitative RT-PCR for SARS-CoV-2***

We performed qRT-PCR for SARS-CoV-2 as previously described. Briefly, proportional amounts of a qualitative internal extraction control (replication-competent avian leukosis virus (ALV) long terminal repeat (LTR) with a splice adaptor (RCAS) (DOI: 10.1128/JCM.41.10.4531-4536.2003) were added to 0.35 mL of inactivated saliva and stool samples (when available), and total RNA extracted using the MagMax Viral Pathogens Kit (kit-supplied Proteinase K replaced with 200µg Ambion ProteinaseK) and the Kingfisher Flex automated extractor (Thermofisher). One step quantitative RT-PCR was performed on RNA extracts using primer/probe sets to detect RCAS (DOI: 10.1128/JCM.41.10.4531-4536.2003), SARS-CoV-2 N, and human RNaseP (Thermofisher TaqPath™ 1-Step RT-qPCR Master Mix, Cat#A15300; RnaseP target from TaqMan 2019nCoV Assay Kit v1, Cat#A47532; N target (Fwd: 5’-GTTTGGTGGACCCTCAGATT-3’, Rev: 5’-CGCAGTATTATTGGGTAAACCTTG-3’, Probe: 5’6-FAM-TAACCAGAATGGAGAACGCAGTGGG-3’BHQ1). RNA was quantified using an in-run standard curve constructed from full genome SARS-CoV-2 RNA (Wuhan-1, Twist Biosciences, Twist Synthetic SARS-CoV-2 RNA Control 2 (MN908947.3) - SKU: 102024) that was concentration-verified by endpoint dilution. Each run contained a positive control (100 copies of heat inactivated SARS-CoV-2 (Isolate USA-WA1/2020, BEI Resources Catalog No. NR-52350) in healthy human pre-COVID specimen), and negative control (healthy human pre-COVID specimen).^[9,10](https://sciwheel.com/work/citation?ids=12208453,13061298&pre=&pre=&suf=&suf=&sa=0,0&dbf=0&dbf=0)^

#### Covariates:

We included the following covariates (clinical variables) in analyses of the primary and secondary outcomes:

- Demographic information: age, sex, race, ethnicity, body mass index, education level, marital status
- Comorbid conditions: hypertension, diabetes, coronary artery disease (CAD), congestive heart failure (CHF), stroke, atrial fibrillation, obstructive sleep apnea (OSA), chronic obstructive pulmonary disease (COPD), asthma, cancer, pulmonary embolism, auto-immune disease, HIV, anemia, total smoking exposure (expressed in pack years).
- Acute COVID-19 severity: inpatient vs. outpatient
- Receipt of antiviral treatment during acute illness
- Time (days) from (most recent) COVID-19 diagnosis from baseline visit assessment
- Number of COVID-19 illnesses
- Number of COVID-19 vaccination doses prior to baseline visit assessment
- Type of COVID-19 vaccination (mRNA vs. viral vector)
- Calendar period of index COVID-19 illness (as proxy for SARS-CoV-2 variant) [https://coronavirus.health.ny.gov/covid-19-variant-data]

#### Statistical procedures

We examined data for distribution and missingness. We constructed descriptive plots for continuous variables (boxplots with individual data points) to examine for outliers and then confirmed data accuracy for outlier observations. For baseline outcome data (symptoms and scales), we performed complete case analyses following extensive efforts to ensure data completeness. We performed non-parametric comparisons for continuous (described as median and interquartile range – IQR) and categorical variables (Kruskal-Wallis, Wilcoxon, and Fisher’s exact tests, respectively) between different groups in primary and secondary outcome analyses (e.g. patients with symptom X present vs. not). We reported the nominal p-values for all tests performed.

We described the proportions of subjects reporting each of the following symptoms at baseline visit and generated descriptive tables with covariate comparisons between subjects with or without each symptom.

We analyzed the following 16 symptoms:

Symptoms

Fever

Chills

Muscle aches

Runny nose

Sore throat

Cough

Dyspnea

Nausea or vomiting

Headache

Abdominal pain

Diarrhea

Loss of taste

Loss of smell

“Brain Fog”

Fatigue

Chest issues

For the numerical clinical scales, we classified subjects into widely-accepted ordinal categories of symptom/functional severity based on the following thresholds:

- GAD7: 0-4 minimal anxiety; 5-9 mild anxiety; 10-14 moderate anxiety; >14 severe anxiety.
- PHQ9: 0-4 no depression; 5-9 mild depression; 10-14 moderate depression; 15-19 moderately severe depression; 20-27: severe depression.
- ISI: 0-7 absence of insomnia; 8-14 sub-threshold insomnia; 15-21 moderate insomnia; 22-28 severe insomnia.
- MoCA-BLIND: <18 abnormal; 18-22 normal.
- MMRC: scale from 0 (breathlessness only with strenuous exercise) to 4 (breathless when dressing). MMRC≥2 was considered clinically significant dyspnea.

To examine for presence of distinct subphenotypes (classes) of symptomatology burden and functional impairment among patients with PASC at the time of their initial assessment, we conducted **unsupervised classification with Latent Class Analysis (LCA)**[^14^](https://sciwheel.com/work/citation?ids=10218043&pre=&suf=&sa=0&dbf=0) for self-reported symptoms at baseline visit.

LCA is a widely used methodology of **Finite Mixture Modeling** (FMM) to determine whether unmeasured or unobserved groups exist within a population.[^15,16^](https://sciwheel.com/work/citation?ids=5746990,8570395&pre=&pre=&suf=&suf=&sa=0,0&dbf=0&dbf=0) The unobserved, or “latent”, groups are inferred from patterns of the observed variables or “indicators” used in the modeling strategy.

LCA models work on the assumption that the observed distribution of the variables is the result of a finite latent (unobserved) mixture of underlying distributions. Using a set of observed indicators, LCA models identify maximum likelihood estimate solutions that best describe these latent classes within which the indicators follow the same distribution. LCA is a probabilistic method of unsupervised clustering because once the model has been fitted, the probability of class membership is estimated for each observation in the cohort and such probabilities can then be used to assign class.[^14^](https://sciwheel.com/work/citation?ids=10218043&pre=&suf=&sa=0&dbf=0)

LCA modeling does not assign individuals to latent classes, but rather generates probabilities for membership in all the identified classes in the model. This is key difference between other clustering methods, such as K-means or hierarchical clustering, which use an arbitrary distance measure to identify clusters. K-means or hierarchical clustering solutions try to find a division in the dataset that maximizes the between-cluster difference and minimizes the within-cluster difference, but solutions can be arbitrary. Thus, other clustering approaches separate the study units into different clusters, whereas LCA estimates the probability that a given study unit belongs to each of the different latent classes. As LCA is model-based, it generates fit statistics, which in turn allows statistical inference when determining the most appropriate number of clusters for a population. In comparison to cluster analyses, LCA is therefore considered a more statistically robust method of clustering. LCA has gained popularity as an unsupervised clustering method in different fields of medicine. For these reasons, we used LCA as the primary statistical approach for defining PASC subphenotypes. In exploratory analyses, we also considered hierarchical clustering if not possible to construct LCA models based on available sample size.

We used an established approach for LCA modeling.[^14^](https://sciwheel.com/work/citation?ids=10218043&pre=&suf=&sa=0&dbf=0) As indicator variables for LCA modeling, we considered

Following data clean-up, we constructed two separate LCA models: i) we used as input variables symptoms reported as present at any time post-COVID-19 (“ever-present”) to examine retrospectively the epidemiologic impact of PASC in the study population (i.e. “epidemiologic clusters” – LCA1), and ii) we used as input variables symptoms “currently-present” at the time of the baseline visit to stratify subjects by active symptomatology (“clinically active clusters” – LCA2). We examined model fit performance by calculating membership probability, entropy, Bayesian Information Criterion, log likelihood ratio and Vuong-Lo-Mendell-Rubin Likelihood Ratio Test between different classes, as well as a clinical relevance criterion of ensuring that each class has at least 5% of observations from the cohort. Following derivation of LCA classes, we then examined for differential symptom burden/type between classes to understand whether the statistical models captured different clinical subphenotypes of the complex PASC syndrome, e.g., distinct subphenotypes of respiratory vs. neurocognitive impairments. We also examined whether the classes were associated with clinical covariate differences.

We used the following statistical software:

- Stata 17.0 (StataCorp. 2021. *Stata Statistical Software: Release 17*. College Station, TX: StataCorp LLC.)
- R version 4.2.0 (2022-04-22), R Core Team (2022). R: A language and environment for statistical computing. R Foundation for Statistical Computing, Vienna, Austria. URL: <https://www.R-project.org/>.
- Mplus 8.8. Los Angeles, CA: Muthén & Muthén.

Figure S1: Heatmap of associations between clinical variables and currently present symptoms. Direction of associations (i.e. odds ratio > or <1) are color coded and the statistically significant effect sizes are shown by numerical odds ratios in the heatmap boxes.

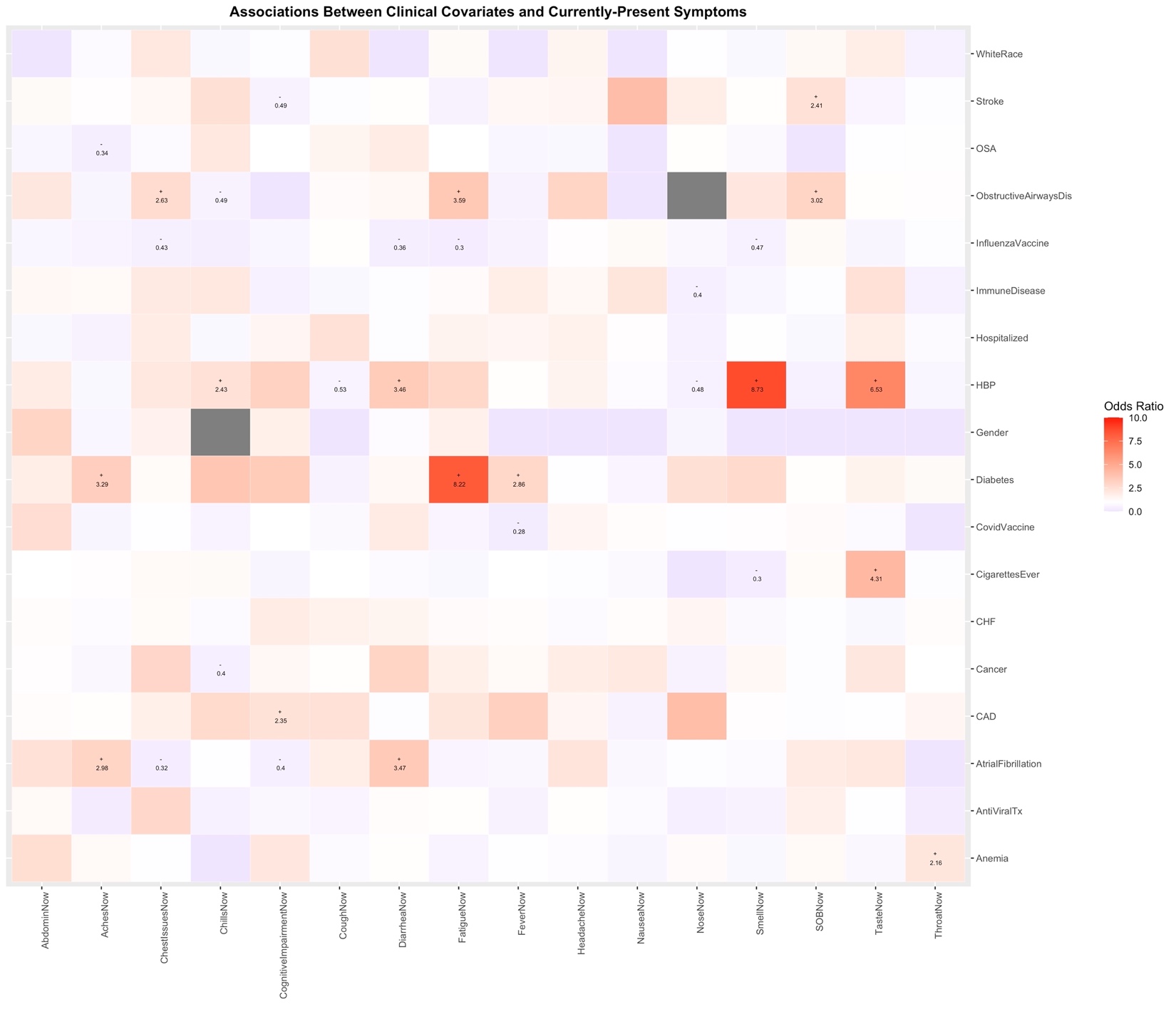

Figure S2: Comparisons in GAD7 scale scores by clinical variables.

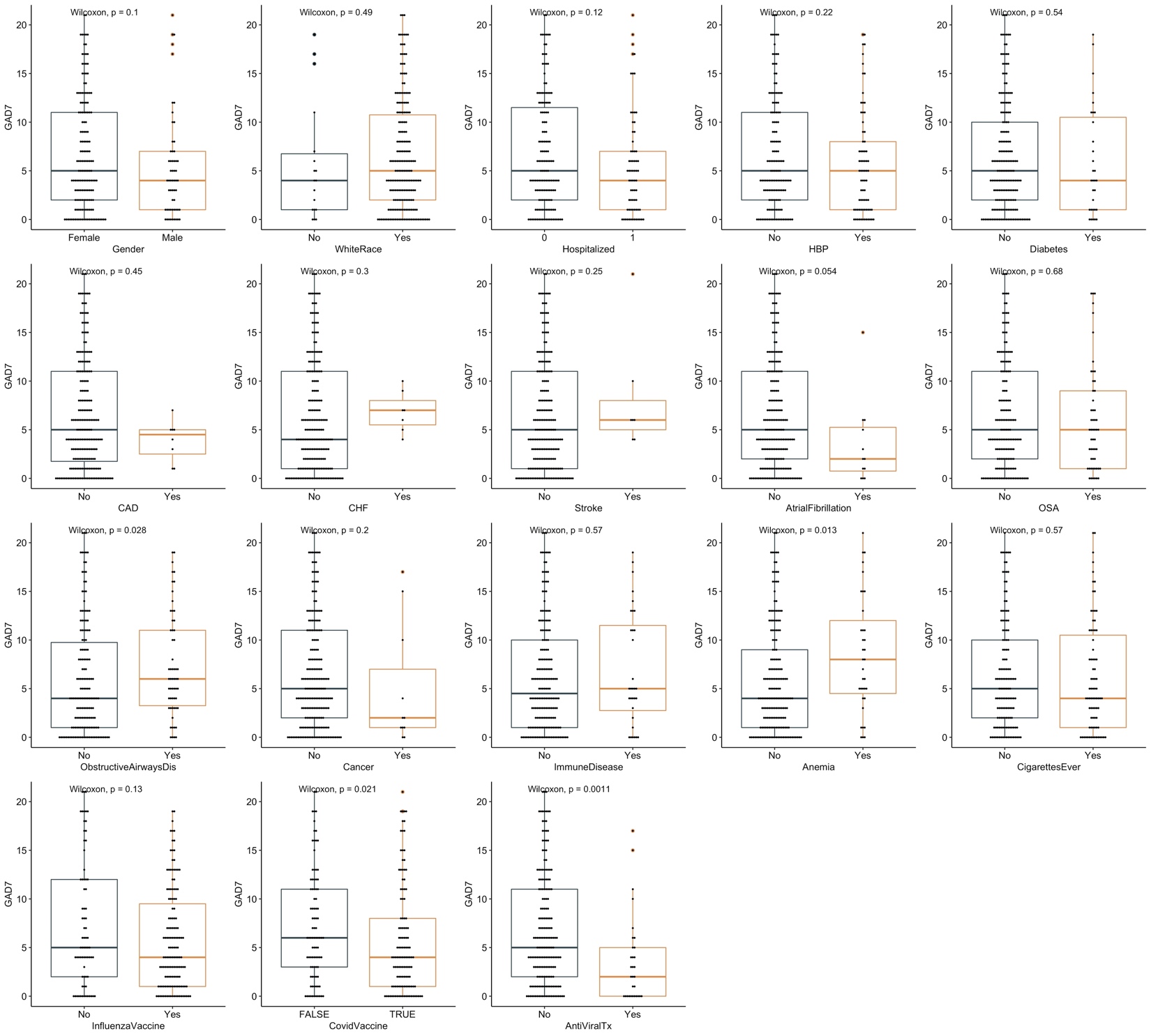

Figure S3: Comparisons in PHQ9 scale scores by clinical variables.

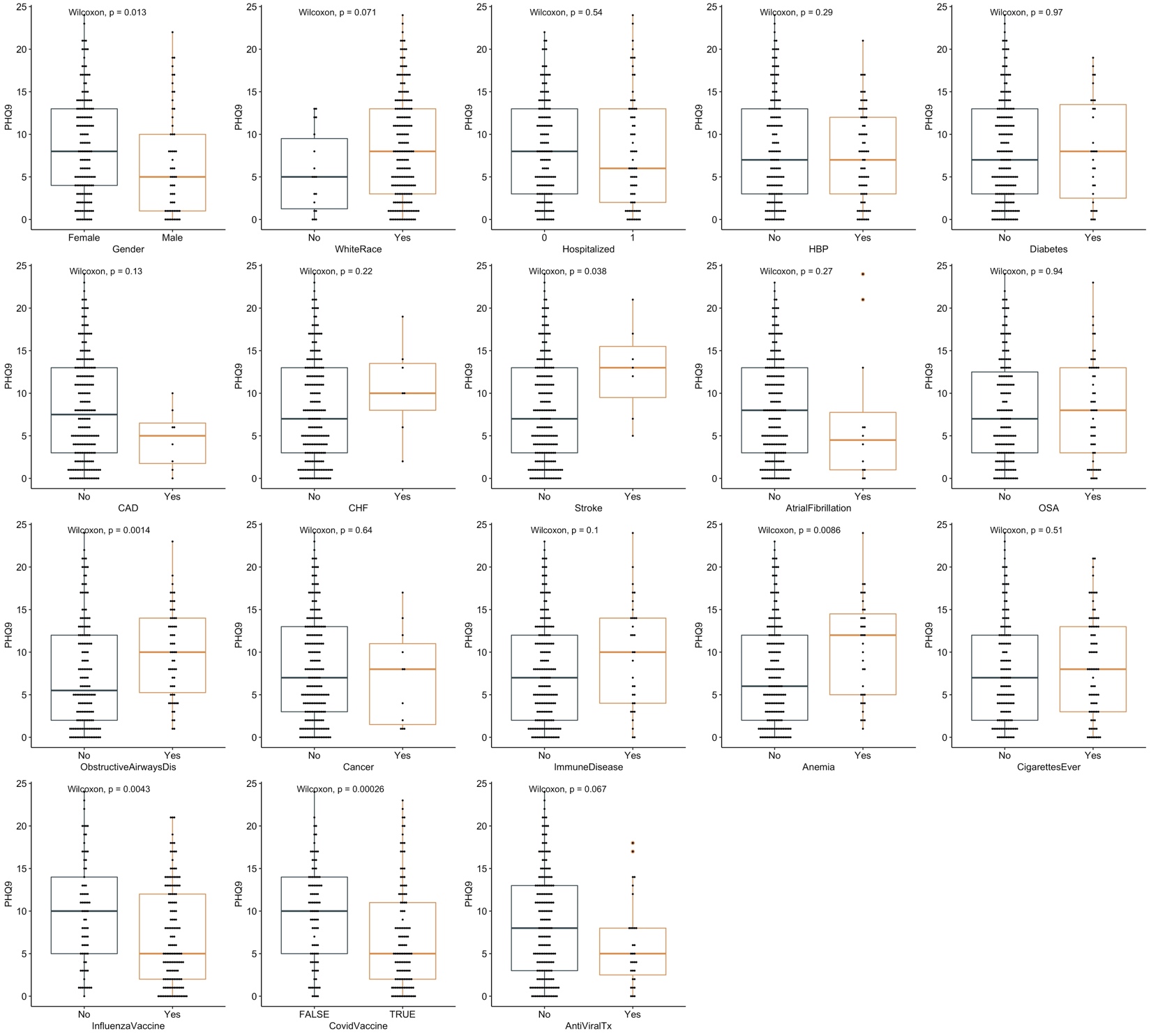

Figure S4: Comparisons in ISI scale scores by clinical variables.

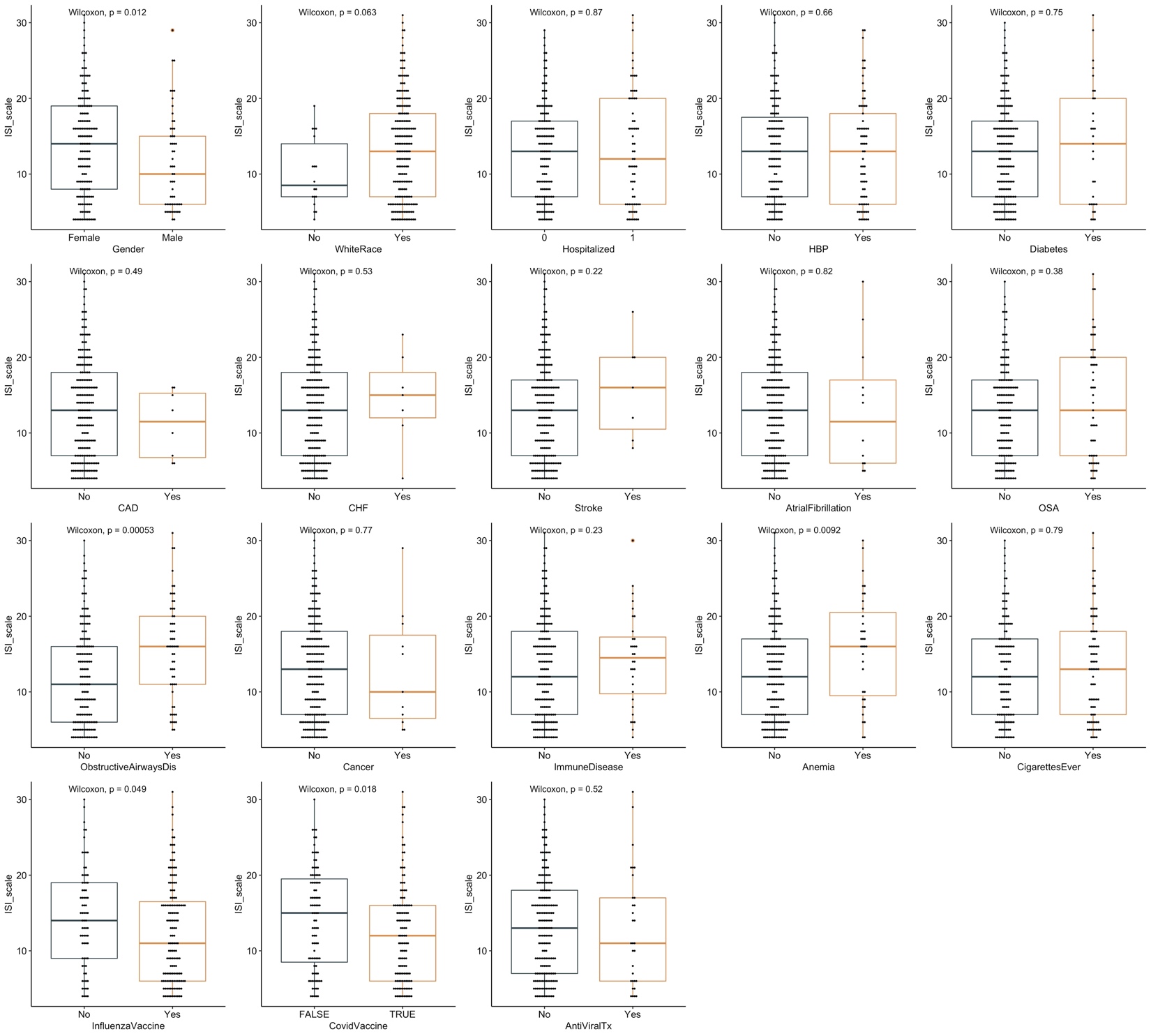

Figure S5: Comparisons in MoCA-BLIND scores by clinical variables.

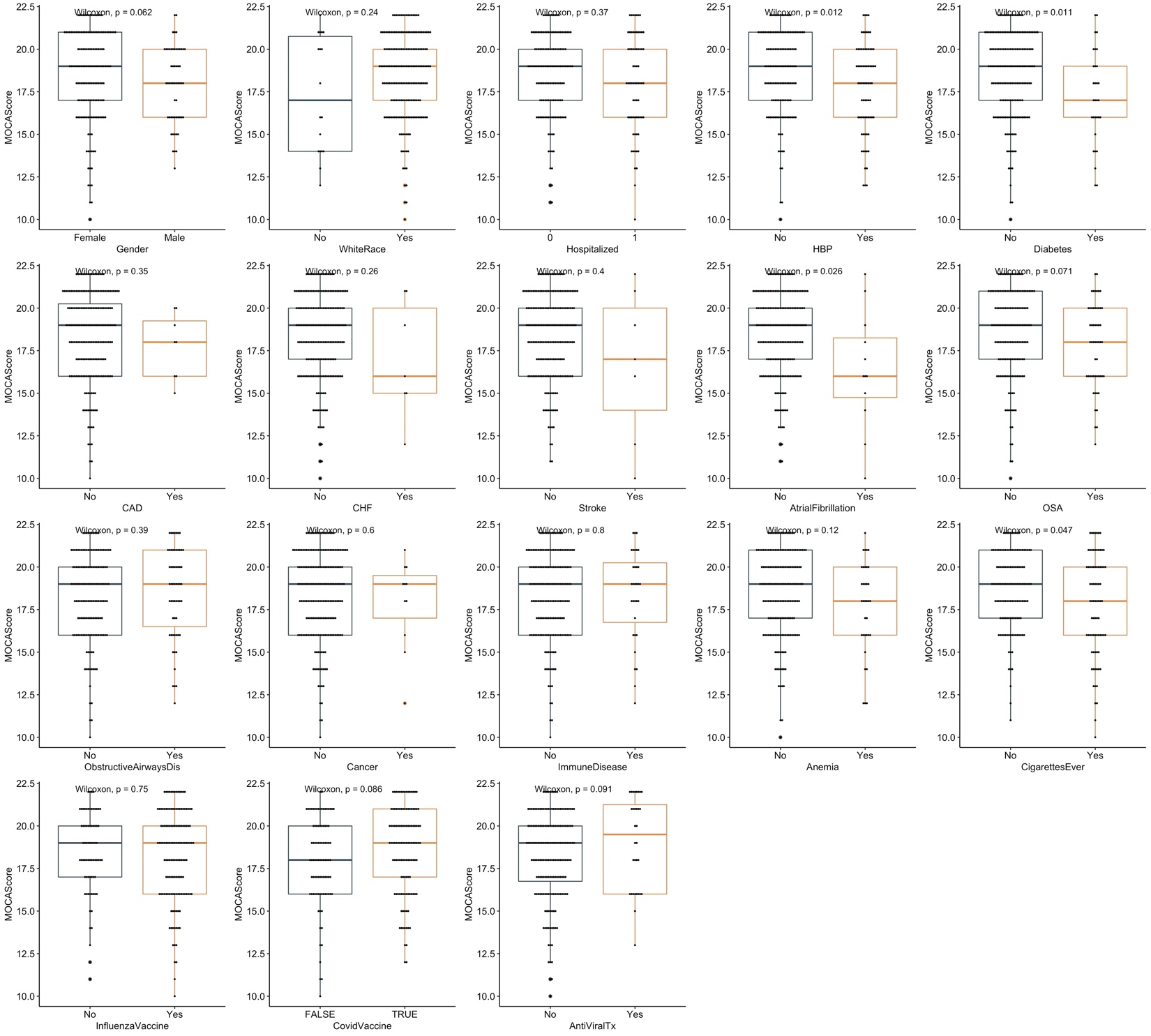

Figure S6: Correlograms between continuous clinical variables and 4 numerical scales.

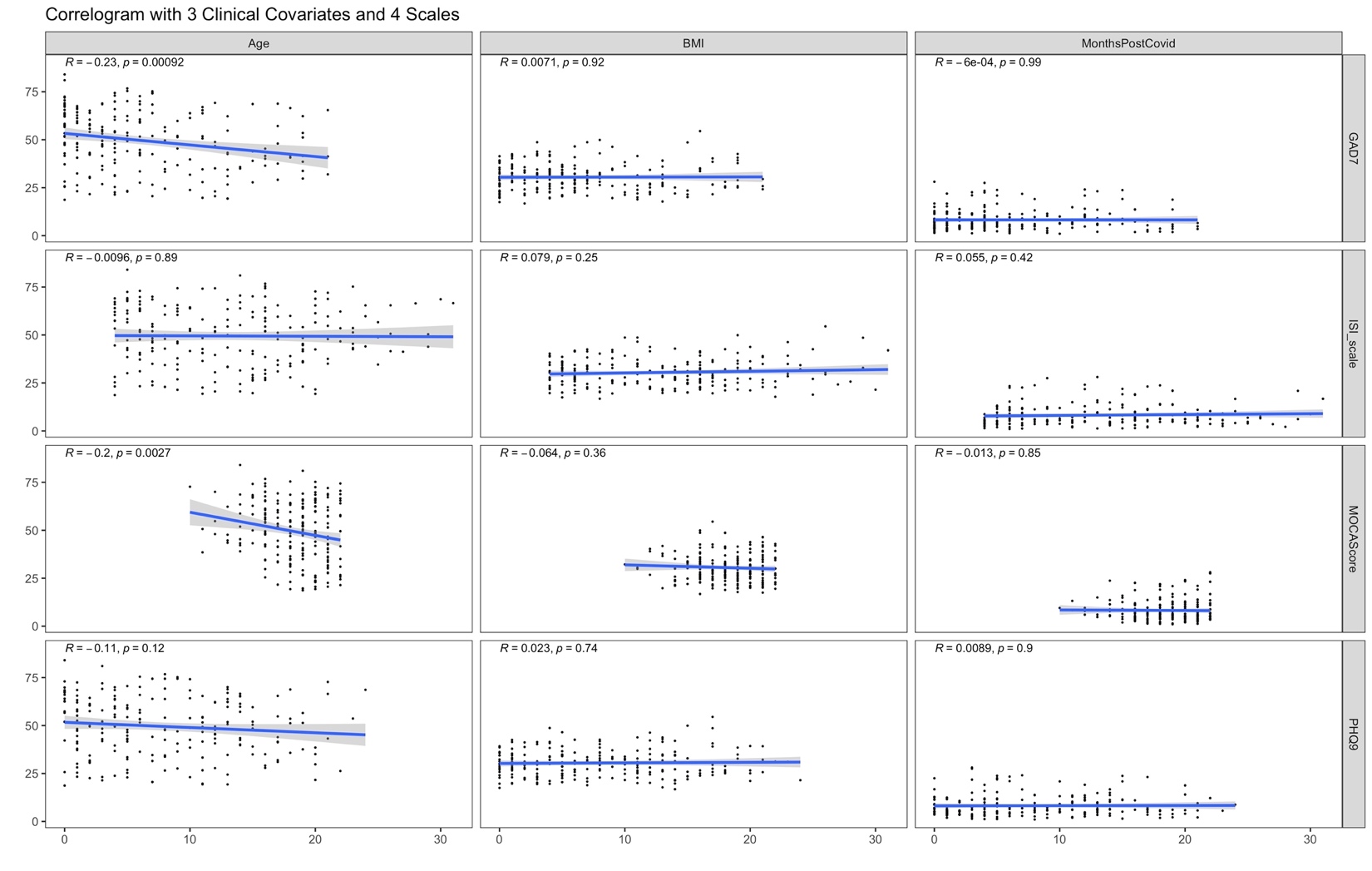

Figure S7: Heatmap of associations between self-reported outcomes with clinical variables. Direction of associations (i.e. odds ratio > or <1) are color coded and the statistically significant effect sizes are shown by numerical odds ratios in the heatmap boxes.

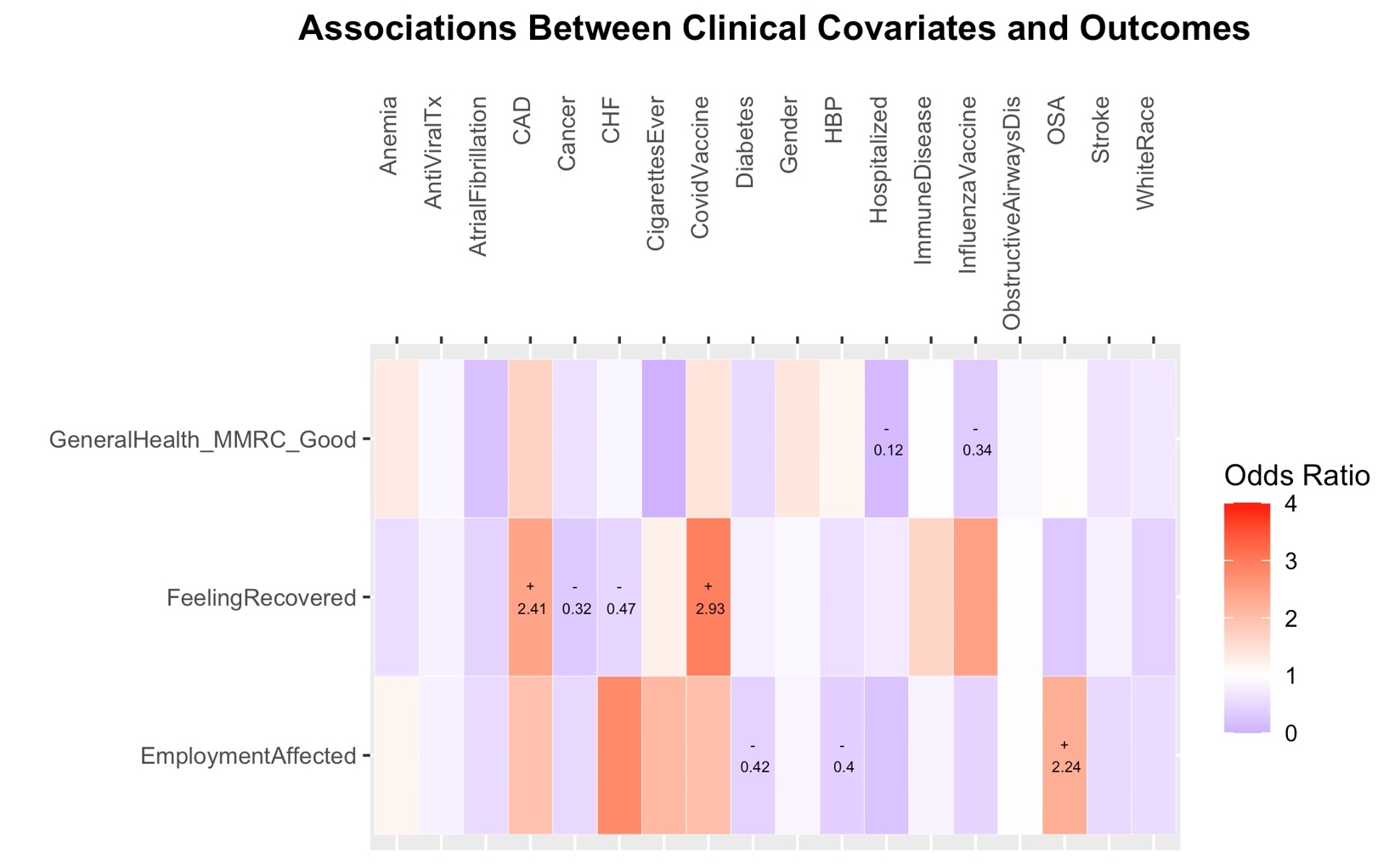

**Figure S8: Distributions of symptoms and numerical scales by the “epidemiologic” subphenotypes of Post-Acute Sequelae of COVID-19 (PASC).** We conducted latent class analysis (LCA) by using the “ever-experienced” 16 symptoms at baseline visit as input variables (“LCA1”). A. Cluster 1 subjects had significantly higher number of “ever-experienced” symptoms compared to cluster 2 subjects, who in turn had much higher number of symptoms compared to cluster 3. B. Stacked bar showing the proportions of presence (“Yes” in orange) vs. absence (“No” in gray) for each of the 16 interviewed symptoms for each of the three clusters. C. Cluster 1 subjects had much higher scores for the numerical scales Generalized Anxiety Scale-7 (GAD7) for anxiety, patient health questionnaire-9 (PHQ9) for depression and insomnia severity index (ISI) for insomnia, but no difference in the Montreal Cognitive Assessment / MoCA-BLIND score for neurocognitive functioning.

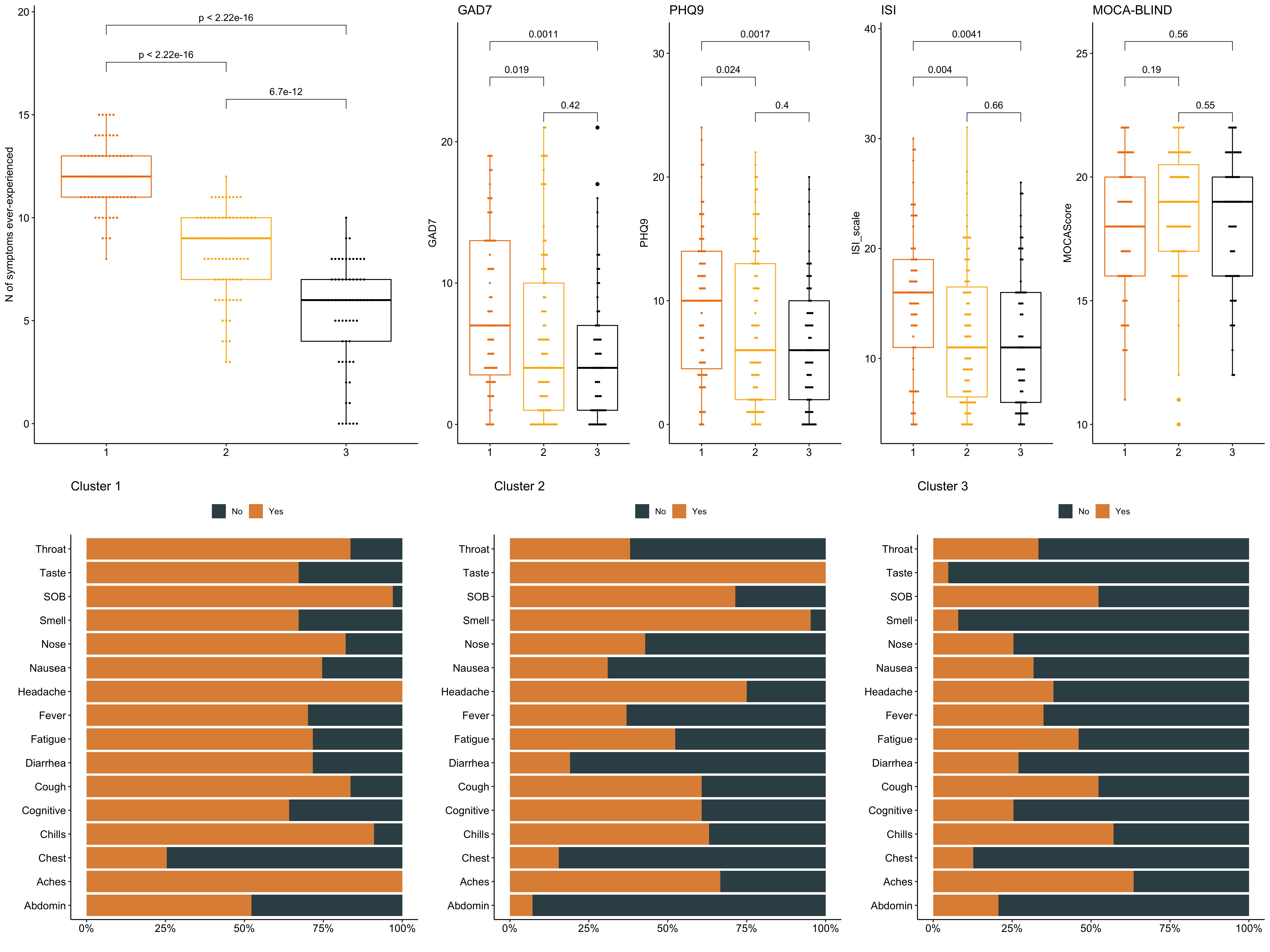

**Figure S9: Cluster 1 and Cluster 2 subjects by LCA-1 had worse outcomes for subjective recovery and impact on employment compared to Cluster 3 (p<0.001).**

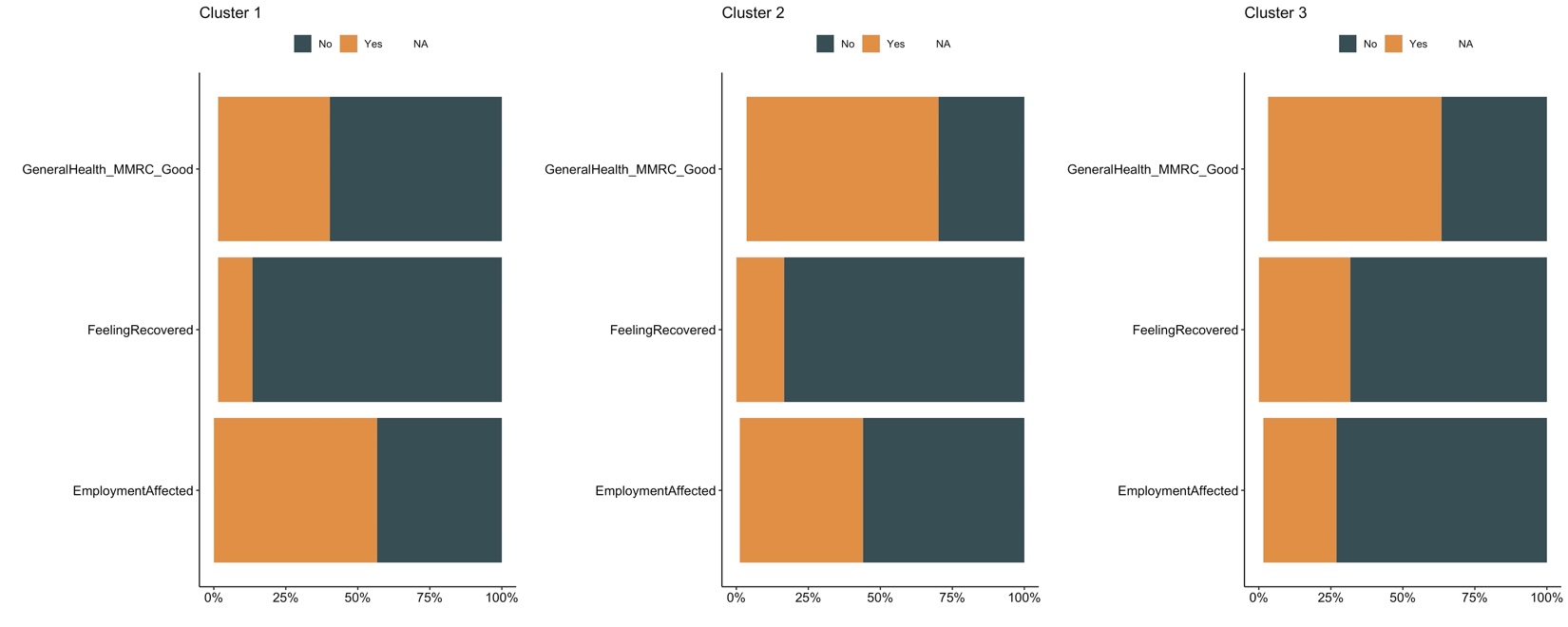

Figure S10: Cluster 1 and Cluster 2 subjects by LCA-2 had worse outcomes for subjective recovery and impact on employment compared to Cluster 3 (p<0.001).

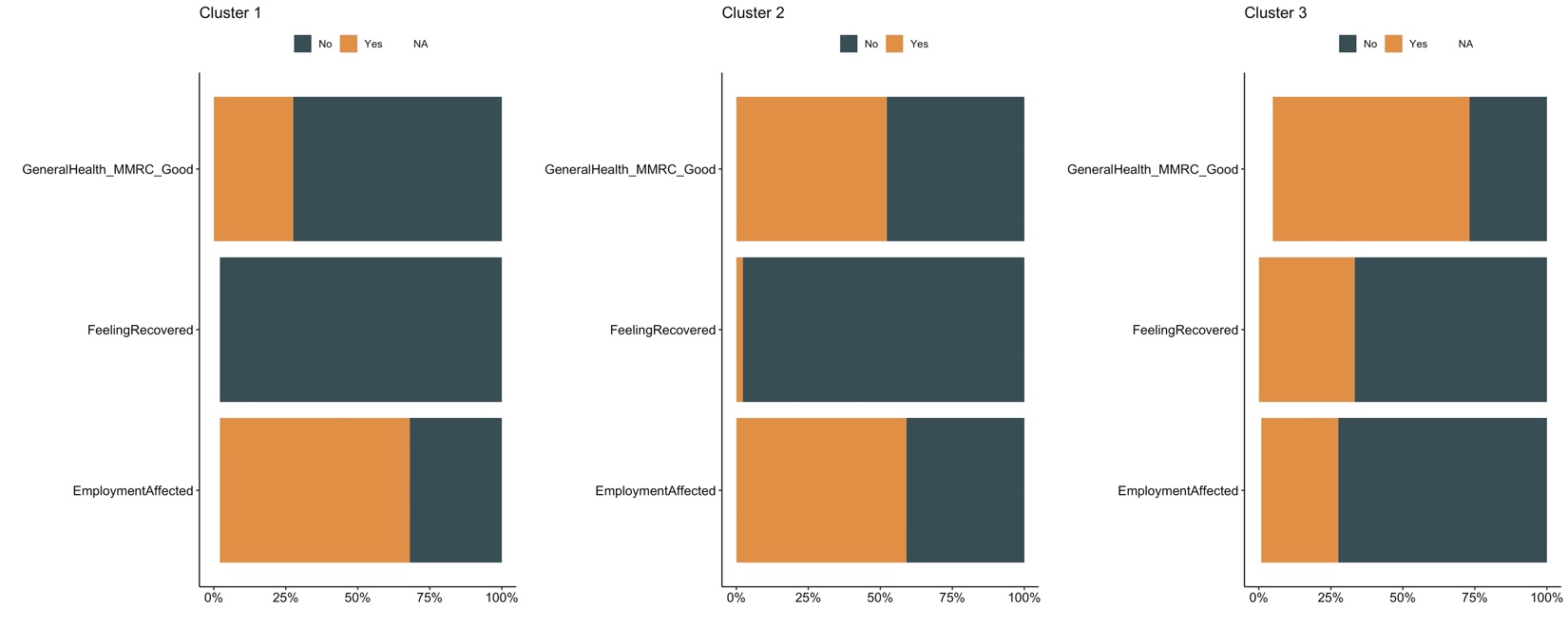

**Table S1: Clinical characteristics of the included subjects with Post Acute Sequelae (PASC) of COVID-19, stratified by referral source.** We present continuous variables as median and interquartile range (IQR) and categorical variables as number (%). We compared continuous variables with Wilcoxon tests and categorical variables with Fisher’s exact tests. We consider p<0.05 as statistically significant.

| **Variable** | **Inpatient Registry** | **Outpatient Study** | **Physician Referral** | **Post Covid Clinic** | **Self-Referral** | **p** |
| --- | --- | --- | --- | --- | --- | --- |
| **Participants** | 42 | 26 | 40 | 68 | 38 |  |
| **Age (median [IQR])** | 63.5 [57.4, 68.9] | 48.2 [42.6, 54.4] | 55.9 [45.8, 65.7] | 48.3 [38.3, 56.3] | 31.3 [24.0, 40.9] | <0.01 |
| **Men (%)** | 14 (33.3) | 7 (26.9) | 11 (27.5) | 22 (32.4) | 3 (7.9) | 0.06 |
| **Whites (%)** | 37 (88.1) | 24 (92.3) | 36 (90.0) | 66 (97.1) | 33 (86.8) | 0.33 |
| **Body Mass Index (BMI) (median [IQR])** | 30.6 [26.6, 35.7] | 30.0 [26.1, 32.8] | 32.5 [26.4, 37.2] | 29.5 [25.1, 33.8] | 26.5 [22.7, 33.8] | 0.07 |
| **Inpatients (%)** | 42 (100.0) | 3 (11.5) | 11 (27.5) | 13 (19.1) | 2 (5.3) | <0.01 |
| **Not graduates of college-level degree (%)** | 25 (59.5) | 9 (34.6) | 23 (57.5) | 30 (44.1) | 14 (36.8) | 0.1 |
| **Hypertension (%)** | 26 (61.9) | 8 (30.8) | 20 (50.0) | 17 (25.0) | 4 (10.5) | <0.01 |
| **Diabetes (%)** | 18 (42.9) | 2 (7.7) | 5 (12.5) | 5 (7.4) | 2 (5.3) | <0.01 |
| **Coronary Artery Disease (%)** | 2 (4.8) | 1 (3.8) | 2 (5.0) | 3 (4.4) | 0 (0.0) | 0.76 |
| **Congestive Heart Failure (%)** | 5 (11.9) | 0 (0.0) | 2 (5.0) | 0 (0.0) | 0 (0.0) | 0.01 |
| **Stroke (%)** | 6 (14.3) | 0 (0.0) | 1 (2.5) | 2 (2.9) | 0 ( 0.0) | 0.01 |
| **Atrial Fibrillation (%)** | 7 (16.7) | 0 (0.0) | 2 (5.0) | 3 (4.4) | 1 (2.6) | 0.03 |
| **OSA (%)** | 18 (42.9) | 4 (15.4) | 11 (27.5) | 17 (25.0) | 4 (10.5) | 0.01 |
| **Obstructive airways disease* (%)** | 15 (35.7) | 7 (26.9) | 9 (22.5) | 19 (27.9) | 10 (26.3) | 0.75 |
| **History of Cancer (%)** | 3 (7.1) | 1 (3.8) | 3 (7.5) | 3 (4.4) | 1 (2.6) | 0.83 |
| **History of Immunosuppression (%)** | 13 (31.0) | 3 (11.5) | 6 (15.0) | 7 (10.3) | 4 (10.5) | 0.04 |
| **Anemia (%)** | 14 (33.3) | 3 (11.5) | 7 (17.5) | 8 (11.8) | 3 (7.9) | 0.01 |
| **Ever smoker (%)** | 21 (50.0) | 8 (30.8) | 16 (40.0) | 24 (35.3) | 11 (28.9) | 0.31 |
| **Vaccinated for Influenza (%)** | 26 (61.9) | 19 (73.1) | 26 (65.0) | 42 (61.8) | 27 (71.1) | 0.76 |
| **Vaccinated for COVID-19 (%)** | 26 (61.9) | 11 (42.3) | 13 (32.5) | 39 (57.4) | 33 (86.8) | <0.01 |
| **No of COVID-19 vaccinations, (median [IQR])** | 2.0 [0.0, 3.0] | 0.0 [0.0, 2.8] | 0.0 [0.0, 2.2] | 1.5 [0.0, 3.0] | 3.0 [2.0, 4.0] | <0.01 |
| **Antiviral Treatment during acute COVID-19 (%)** | 23 (54.8) | 0 (0.0) | 1 (2.5) | 5 (7.4) | 4 (10.5) | <0.01 |
| **Probable SARS-CoV-2 variant** |  |  |  |  |  | <0.01 |
| **Alpha** | 11 (26.2) | 18 (69.2) | 19 (47.5) | 22 (32.4) | 3 (7.9) |  |
| **Delta** | 0 (0.0) | 1 (3.8) | 3 (7.5) | 27 (39.7) | 6 (15.8) |  |
| **Omicron** | 0 (0.0) | 0 (0.0) | 1 (2.5) | 5 (7.4) | 25 (65.8) |  |
| **Wild Type** | 31 (73.8) | 7 (26.9) | 17 (42.5) | 14 (20.6) | 4 (10.5) |  |
| **Days post-acute COVID-19, (median [IQR])** | 148.5 [124.5, 197.5] | 185.5 [163.0, 220.8] | 219.5 [153.5, 266.2] | 241.5 [141.2, 404.5] | 300.0 [167.0, 370.2] | <0.01 |

**Table S2: Clinical characteristics by LCA-1 clusters.**

| Variable | 1 | 2 | 3 | P Value |
| --- | --- | --- | --- | --- |
| Participants | 67 | 84 | 63 |  |
| Age (median [IQR]) | 49.5 [39.6, 58.1] | 49.6 [40.2, 60.5] | 54.1 [36.1, 66.0] | 0.35 |
| Men (%) | 11 (16.4) | 25 (29.8) | 21 (33.3) | 0.07 |
| Whites (%) | 60 (89.6) | 81 (96.4) | 55 (87.3) | 0.11 |
| Body Mass Index (BMI) (median [IQR]) | 29.8 [25.4, 33.7] | 29.8 [25.1, 36.0] | 30.1 [25.1, 34.4] | 0.85 |
| Inpatients (%) | 17 (25.4) | 25 (29.8) | 29 (46.0) | 0.03 |
| Not graduates of college-level degree (%) | 35 (52.2) | 35 (41.7) | 31 (49.2) | 0.4 |
| Hypertension (%) | 21 (31.3) | 27 (32.1) | 27 (42.9) | 0.3 |
| Diabetes (%) | 9 (13.4) | 10 (11.9) | 13 (20.6) | 0.31 |
| Coronary Artery Disease (%) | 3 (4.5) | 2 (2.4) | 3 (4.8) | 0.7 |
| Congestive Heart Failure (%) | 2 (3.0) | 2 (2.4) | 3 (4.8) | 0.72 |
| Stroke (%) | 1 (1.5) | 4 (4.8) | 4 (6.3) | 0.37 |
| Atrial Fibrillation (%) | 3 (4.5) | 4 (4.8) | 6 (9.5) | 0.39 |
| OSA (%) | 15 (22.4) | 23 (27.4) | 16 (25.4) | 0.78 |
| Obstructive airways disease* (%) | 28 (41.8) | 18 (21.4) | 14 (22.2) | 0.01 |
| History of Cancer (%) | 6 (9.0) | 2 (2.4) | 3 (4.8) | 0.19 |
| History of Immunosuppression (%) | 12 (17.9) | 11 (13.1) | 10 (15.9) | 0.71 |
| Anemia (%) | 14 (20.9) | 12 (14.3) | 9 (14.3) | 0.48 |
| Ever smoker (%) | 27 (40.3) | 27 (32.1) | 26 (41.3) | 0.44 |
| Vaccinated for Influenza (%) | 47 (70.1) | 53 (63.1) | 40 (63.5) | 0.62 |
| Vaccinated for COVID-19 (%) | 39 (58.2) | 45 (53.6) | 38 (60.3) | 0.7 |
| No of COVID-19 vaccinations, median [IQR | 2.0 [0.0, 3.0] | 1.5 [0.0, 3.0] | 2.0 [0.0, 3.0] | 0.53 |
| Antiviral Treatment during acute COVID-19 (%) | 7 (10.4) | 13 (15.5) | 13 (20.6) | 0.27 |
| Probable SARS-CoV-2 variant |  |  |  | <0.01 |
| Alpha | 17 (25.4) | 39 (46.4) | 17 (27.0) |  |
| Delta | 16 (23.9) | 17 (20.2) | 4 (6.3) |  |
| Omicron | 13 (19.4) | 8 (9.5) | 10 (15.9) |  |
| WildType | 21 (31.3) | 20 (23.8) | 32 ( 50.8) |  |
| Days post acute COVID-19, (median [IQR]) | 250.0 [154.0, 390.5] | 180.0 [132.0, 290.8] | 190.0 [144.0, 282.0] | 0.09 |

**Table S3: Clinical characteristics by LCA-2 clusters**

| Variable | 1 | 2 | 3 | P Value |
| --- | --- | --- | --- | --- |
| Participants | 47 | 44 | 123 |  |
| Age (median [IQR]) | 47.3 [39.1, 54.4] | 54.4 [45.6, 65.5] | 49.8 [34.9, 62.6] | 0.06 |
| Men (%) | 9 (19.1) | 11 (25.0) | 37 (30.1) | 0.34 |
| Whites (%) | 45 (95.7) | 42 (95.5) | 109 (88.6) | 0.19 |
| Body Mass Index (BMI) (median [IQR]) | 30.2 [25.1, 35.8] | 30.0 [24.7, 34.9] | 30.0 [25.4, 34.4] | 0.77 |
| Inpatients (%) | 10 (21.3) | 14 (31.8) | 47 (38.2) | 0.11 |
| Not graduates of college-level degree (%) | 26 (55.3) | 21 (47.7) | 54 (43.9) | 0.41 |
| Hypertension (%) | 20 (42.6) | 12 (27.3) | 43 (35.0) | 0.31 |
| Diabetes (%) | 10 (21.3) | 4 (9.1) | 18 (14.6) | 0.26 |
| Coronary Artery Disease (%) | 1 (2.1) | 3 (6.8) | 4 (3.3) | 0.45 |
| Congestive Heart Failure (%) | 1 (2.1) | 1 (2.3) | 5 (4.1) | 0.75 |
| Stroke (%) | 2 (4.3) | 2 (4.5) | 5 (4.1) | 0.99 |
| Atrial Fibrillation (%) | 3 (6.4) | 2 (4.5) | 8 (6.5) | 0.89 |
| OSA (%) | 14 (29.8) | 12 (27.3) | 28 (22.8) | 0.6 |
| Obstructive airways disease* (%) | 18 (38.3) | 12 (27.3) | 30 ( 24.4) | 0.19 |
| History of Cancer (%) | 5 (10.6) | 0 (0.0) | 6 (4.9) | 0.07 |
| History of Immunosuppression (%) | 11 (23.4) | 5 (11.4) | 17 (13.8) | 0.21 |
| Anemia (%) | 11 (23.4) | 4 (9.1) | 20 (16.3) | 0.18 |
| Ever smoker (%) | 18 (38.3) | 20 (45.5) | 42 (34.1) | 0.41 |
| Vaccinated for Influenza (%) | 26 (55.3) | 31 (70.5) | 83 (67.5) | 0.24 |
| Vaccinated for COVID-19 (%) | 20 (42.6) | 25 (56.8) | 77 (62.6) | 0.06 |
| No of COVID-19 vaccinations, (median [IQR]) | 0.0 [0.0, 2.0] | 2.0 [0.0, 3.0] | 2.0 [0.0, 3.0] | 0.01 |
| Antiviral Treatment during acute COVID-19 (%) | 4 (8.5) | 6 (13.6) | 23 (18.7) | 0.24 |
| Probable SARS-CoV-2 variant |  |  |  | 0.3 |
| Alpha | 15 (31.9) | 17 (38.6) | 41 (33.3) |  |
| Delta | 11 (23.4) | 9 (20.5) | 17 (13.8) |  |
| Omicron | 6 (12.8) | 2 (4.5) | 23 (18.7) |  |
| WildType | 15 (31.9) | 16 (36.4) | 42 (34.1) |  |
| Days post acute COVID-19, (median [IQR]) | 190.0 [145.5, 344.5] | 246.0 [163.5, 388.8] | 184.0 [129.0, 290.0] | 0.17 |

**Table S4: LASSO model predictor of LCA-2 clusters.**

| **Variable** | **Co-efficient** |
| --- | --- |
| Intercept | -1.489 |
| BMI | 0.053 |
| Hospitalized | 0.491 |
| COVID vaccination | 0.630 |
| Education (Not graduates of college degree) | -0.0241 |
| Anemia | -0.696 |
| History of Autoimmune disease | -0.264 |
| Infection with the delta variant | 0.143 |

**Table S5: Clinical characteristics for subjects with positive vs. negative viral RNA load in saliva samples**

| Variable | Negative | Positive | P Value |
| --- | --- | --- | --- |
| Participants | 97 | 6 |  |
| Age (median [IQR]) | 56.5 [43.9, 65.4] | 66.1 [61.4, 68.5] | 0.2 |
| Men (%) | 33 (34.0) | 2 (33.3) | 1 |
| Whites (%) | 89 (91.8) | 6 (100.0) | 1 |
| Body Mass Index (BMI) (median [IQR]) | 30.5 [26.6, 34.7] | 29.4 [21.6, 34.2] | 0.46 |
| Inpatients (%) | 45 (46.4) | 4 (66.7) | 0.59 |
| Not graduates of college-level degree (%) | 43 (44.3) | 3 (50.0) | 1 |
| Hypertension (%) | 42 (43.3) | 3 (50.0) | 1 |
| Diabetes (%) | 18 (18.6) | 1 (16.7) | 1 |
| Coronary Artery Disease (%) | 5 (5.2) | 2 (33.3) | 0.07 |
| Congestive Heart Failure (%) | 4 (4.1) | 0 (0.0) | 1 |
| Stroke (%) | 6 (6.2) | 1 (16.7) | 0.88 |
| Atrial Fibrillation (%) | 7 (7.2) | 1 (16.7) | 0.96 |
| OSA (%) | 25 (25.8) | 1 (16.7) | 0.99 |
| Obstructive airways disease* (%) | 26 (26.8) | 3 (50.0) | 0.45 |
| History of Cancer (%) | 5 (5.2) | 0 (0.0) | 1 |
| History of Immunosuppression (%) | 18 (18.6) | 1 ( 16.7) | 1 |
| Anemia (%) | 16 (16.5) | 3 (50.0) | 0.13 |
| Ever smoker (%) | 39 (40.2) | 3 (50.0) | 0.96 |
| Vaccinated for Influenza (%) | 61 (62.9) | 5 (83.3) | 0.57 |
| Vaccinated for COVID-19 (%) | 47 (48.5) | 5 (83.3) | 0.22 |
| No of COVID-19 vaccinations, (median [IQR]) | 0.0 [0.0, 3.0] | 3.0 [2.2, 3.8] | 0.02 |
| Antiviral Treatment during acute COVID-19 (%) | 17 (17.5) | 4 (66.7) | 0.02 |
| Probable SARS-CoV-2 variant |  |  |  |
| Alpha | 38 (39.2) | 3 (50.0) |  |
| Delta | 9 (9.3) | 1 (16.7) |  |
| Omicron | 0 0.0) | 0 (0.0) |  |
| Wild Type | 50 (51.5) | 2 (33.3) |  |
| Days post-acute COVID-19, median [IQR] | 188.0 [145.0, 271.0] | 133.0 [58.8, 174.2] | 0.05 |

**Table S6: self-reported symptoms for subjects with positive vs. negative viral RNA load in saliva samples**

| **Saliva Virus Positive** | | | |
| --- | --- | --- | --- |
| **Renamed Columns** | **No** | **Yes** | **p** |
| **Participants** | 97 | 6 |  |
| **Fever ever** | 39 (40.2) | 1 (16.7) | 0.47 |
| **Fever at the time of questionnaire** | 3 (7.3) | 0 (0.0) | 1 |
| **Feverish ever** | 64 (66.0) | 1 (16.7) | 0.05 |
| **Feverish at the time of questionnaire** | 9 (14.1) | 0 (0.0) | 1 |
| **Chills ever** | 70 (72.2) | 4 (66.7) | 1 |
| **Chills at the time of questionnaire** | 8 (11.6) | 1 (25.0) | 0.99 |
| **Muscle Aches ever** | 74 (76.3) | 2 (33.3) | 0.07 |
| **Muscle Aches at the time of questionnaire** | 32 (43.8) | 2 (100.0) | 0.39 |
| **Runny nose ever** | 37 (38.1) | 1 (16.7) | 0.53 |
| **Runny nose at the time of questionnaire** | 14 (34.1) | 0 (0.0) | 1 |
| **Sore throat ever** | 46 (47.4) | 1 (16.7) | 0.3 |
| **Sore throat at the time of questionnaire** | 13 (26.5) | 0 (0.0) | 1 |
| **Cough ever** | 56 (57.7) | 2 (33.3) | 0.46 |
| **Cough at the time of questionnaire** | 23 (39.7) | 1 (50.0) | 1 |
| **Shortness of breath ever** | 75 (77.3) | 2 (33.3) | 0.05 |
| **Shortness of breath at the time of questionnaire** | 41 (54.7) | 2 (100.0) | 0.58 |
| **Nausea or vomiting ever** | 43 (44.3) | 1 (16.7) | 0.37 |
| **Nausea or vomiting at the time of questionnaire** | 13 (29.5) | 0 (0.0) | 1 |
| **Headache ever** | 65 (67.0) | 2 (33.3) | 0.22 |
| **Headache at the time of questionnaire** | 24 (36.4) | 2 (100.0) | 0.28 |
| **Abdominal pain ever** | 23 (23.7) | 0 (0.0) | 0.4 |
| **Abdominal pain at the time of questionnaire** | 14 (51.9) | NA | NA |
| **Diarrhea ever** | 40 (41.2) | 1 (16.7) | 0.45 |
| **Diarrhea at the time of questionnaire** | 16 (38.1) | 0 (0.0) | 1 |
| **Loss of taste ever** | 61 (62.9) | 4 (66.7) | 1 |
| **Loss of taste at the time of questionnaire** | 22 (34.9) | 2 (50.0) | 0.94 |
| **Loss of smell ever** | 59 (60.8) | 4 (66.7) | 1 |
| **Loss of smell at the time of questionnaire** | 26 (42.6) | 2 (50.0) | 1 |
| **Cognitive impairment ever** | 45 (46.4) | 1 (16.7) | 0.32 |
| **Cognitive impairment at the time of questionnaire** | 40 (88.9) | 1 (100.0) | 1 |
| **Fatigue ever** | 39 (40.2) | 3 (50.0) | 0.96 |
| **Fatigue at the time of questionnaire** | 31 (79.5) | 2 (66.7) | 1 |
| **Chest complaints ever** | 4 (4.1) | 0 (0.0) | 1 |
| **Chest complaints at the time of questionnaire** | 3 (75.0) | 0 (NaN) | NaN |
| **Other symptom ever** | 46 (51.1) | 2 (33.3) | 0.67 |
| **Other symptoms at the time of questionnaire** | 35 (74.5) | 1 (50.0) | 1 |
| **ISI Sleep Hours (median [IQR])** | 6.0 [6.0, 7.2] | 6.0 [5.0, 7.0] | 0.4 |
| **GAD7 (median [IQR])** | 4.0 [1.0, 11.0] | 5.5 [3.5, 9.0] | 0.92 |
| **PHQ 9 (median [IQR])** | 8.0 [2.0, 13.0] | 6.5 [4.2, 8.0] | 0.43 |
| **ISI scale (median [IQR])** | 12.0 [6.0, 19.0] | 7.5 [6.2, 13.2] | 0.34 |
| **MOCA Score (median [IQR])** | 18.0 [16.0, 20.5] | 18.5 [16.5, 19.0] | 0.89 |
| **MMRC (median [IQR])** | 1.0 [0.0, 3.0] | 2.5 [1.2, 3.0] | 0.29 |
| **Endurance Exercise Days (median [IQR])** | 1.5 [0.0, 4.0] | 0.0 [0.0, 1.5] | 0.22 |
| **Endurance Exercise Mins (median [IQR])** | 20.0 [0.0, 36.2] | 0.0 [0.0, 11.2] | 0.07 |
| **Fever Days (median [IQR])** | 7.0 [3.0, 14.0] | 2.0 [2.0, 2.0] | 0.21 |
| **Feverish Days (median [IQR])** | 7.0 [4.0, 15.8] | 2.0 [2.0, 2.0] | 0.13 |
| **Chills Days (median [IQR])** | 7.0 [3.0, 14.0] | 4.5 [3.5, 10.0] | 0.59 |
| **Muscle aches Days (median [IQR])** | 20.5 [7.0, 153.8] | 20.0 [15.0, 25.0] | 0.86 |
| **Runny Nose Days (median [IQR])** | 21.0 [8.0, 90.0] | 2.0 [2.0, 2.0] | 0.1 |
| **Throat Days (median [IQR])** | 10.0 [4.2, 47.0] | 4.0 [4.0, 4.0] | 0.3 |
| **Cough Days (median [IQR])** | 30.0 [13.5, 150.2] | 15.0 [12.5, 17.5] | 0.35 |
| **Shortness of breath Days (median [IQR])** | 60.0 [14.0, 213.5] | 34.0 [32.0, 36.0] | 0.63 |
| **Nausea or vomiting Days (median [IQR])** | 14.0 [8.5, 71.5] | 4.0 [4.0, 4.0] | 0.18 |
| **Headache Days (median [IQR])** | 21.0 [10.0, 106.0] | 8.5 [5.2, 11.8] | 0.22 |
| **Abdominal Pain Days (median [IQR])** | 78.0 [12.0, 237.5] | NA [NA, NA] | NA |
| **Diarrhea Days (median [IQR])** | 12.0 [5.8, 68.2] | 3.0 [3.0, 3.0] | 0.16 |
| **Loss of Taste Days (median [IQR])** | 30.0 [14.0, 150.0] | 10.5 [5.8, 18.0] | 0.06 |
| **Loss of Smell Days (median [IQR])** | 60.0 [21.0, 157.0] | 10.5 [5.8, 18.0] | 0.03 |
| **Cognitive Impairment Days (median [IQR])** | 148.0 [65.0, 271.0] | 38.0 [38.0, 38.0] | 0.19 |
| **Fatigue Days (median [IQR])** | 212.0 [60.0, 296.0] | 30.0 [17.0, 34.0] | 0.02 |
| **Chest Complaint Days (median [IQR])** | 158.0 [52.5, 287.0] | NA [NA, NA] | NA |
